# Twenty years review of probiotic meta-analyses articles: Effects on disease prevention and treatment

**DOI:** 10.1101/2021.08.23.21262411

**Authors:** Kajal Farahmandi, Sadegh Sulaimany, Kambiz Kalhor

## Abstract

The study of the probiotic effect in the prevention or treatment of diseases has long attracted the attention of many researchers. Here, we collected close to 300 meta-analysis articles for 20 years, investigating the effect of probiotics in the prevention and treatment of diseases. The goal of this study is to provide an overview of all meta-analysis articles of the effects of probiotics on various human diseases. For this purpose, different online databases, Pubmed, ScienceDirect, and Google Scholar, were searched with the keywords “probiotics” + “disease” + “meta-analysis” in the title, abstract, and keywords. Papers studied and categorized and investigated in order to present valuable insights for researchers in the field. Some of main categories are based on publication year, publishing journals, gender, age, effect type, disease type, contradicting reports and etc. According to the results, most meta-analyses indicated probiotics were 79% effective in preventing or treating the diseases. Some articles have also reported no positive effects, but there is not any paper in our study confirming the detrimental influence of probiotic effect on human health. For the future works, Cochrane reviews, meta-analysis including dozens of articles (as e.g. for NEC and AAD) may be investigated.

## Introduction

Close to 300 meta-analyses investigating the effect of probiotics in the prevention and treatment of diseases have been published from 2000 to 2020, indicating the importance of probiotics in such areas. Health professionals have confirmed the beneficial effects of probiotics and probiotic foods on human health. Continuous use of probiotics may have a significant impact on reducing the incidence of various diseases, especially for high-risk populations such as hospitalized children, children who are not breast-fed, or live in deprived areas (1).

Recent scientific research on the properties and function of living microorganisms in food and dietary supplements indicates that probiotics may play an essential role in immune, gastrointestinal, and respiratory function and can have considerable impact on reducing the occurrence of infectious diseases especially in children and other at-risk individuals (2). According to Amara and Shibl, probiotics can be useful not only in supporting health or controlling pathogenic infections but also in treating and managing diseases (3).

Numerous meta-analyses help to understand the potential of probiotics on disease. Given the sheer volume of research and discrepancies between study results, combining the results of such studies maybe necessary to obtain a comprehensive overview of the field (4). The number of published meta-analyses on the effect of probiotics in the prevention and treatment of diseases has increased dramatically, as is depicted in supplemental Figure S1. Supplemental Table S1 also provides a list of journals that have published the largest number of meta-analysis articles in this field.

To the best of our knowledge, although a large number of meta-analyses have been published on the effects of probiotics in the prevention and treatment of diseases, no comprehensive overview has been provided so far. Therefore, a review of these articles can be useful and practical in several areas, among which the most important are the researchers’ consensus on whether or not probiotics are useful in the prevention and/or treatment of various diseases and their efficacy. Paradoxes and conflicting reports will also become apparent. Further, the most prevalent diseases in the research and accuracy of reports are revealed, which can help identify future potential areas and research trends.

The goal of this study is to provide an overview of all meta-analysis articles that have investigated the effects of probiotics on various human diseases. We intended to precisely determine the role of probiotics in the prevention and treatment of diseases based on scrutinizing the related meta-analyses. To this end, this study has attempted to provide a comprehensive overview on general and details of existing meta-analyses to determine the effect of probiotics in the prevention and treatment of various diseases and the number of published meta-analysis articles for each disease, as well as categorizing based on disease, age, gender, year of publication, and inconsistent or ambiguous results. As with most meta-analyses, they combine the findings of studies with different probiotic strains or strain combinations. Although health benefits are strain specific, a synthesis of meta-analyses gives indications on what health areas are most promising for investigating and using probiotics.

## Material and methods

For an extensive review of whether or not probiotics are useful in the prevention and treatment of various diseases, including: respiratory, gastrointestinal, mental, skin, infectious, etc., all published meta-analyses articles on probiotics, and disease should be reviewed. Different online databases such as Pubmed, ScienceDirect, and Google Scholar were searched. The search was performed with the keywords “probiotics” + “disease” + “meta-analysis” in the title, abstract, or keywords.

The first article in this field was published in 2000, and the last review we covered in this research was in October 2019. A total of 294 related articles was found. In this study, the classification of diseases was performed according to the International Classification of Diseases system (ICD-10 code). No meta-analysis articles that examined the probiotic effect of disease prevention or treatment, as well as master’s and doctoral theses, descriptive review articles, and reference books were excluded.

## Results and Discussion

Here, we categorize the 294 articles included in our study and present them based on various features and perspectives to help find new ideas and outlooks (Figure 1). These features include the distribution of articles by gender, age, the effectiveness of probiotics for disease treatment, and the distribution of articles by disease type. An adapted PRISMA flow diagram shows the process followed to select the papers used in this research (Figure 1). In total, 26996 publications were reviewed. Finally, 294 were deemed suitable for incorporation, based on the inclusion and exclusion criteria in the analysis. Among the 294 study, some of were related to human health apart from diseases, and some investigated more than one disease or condition in one report. The net number of articles directly related to the diseases was 283.

**Figure 1.**
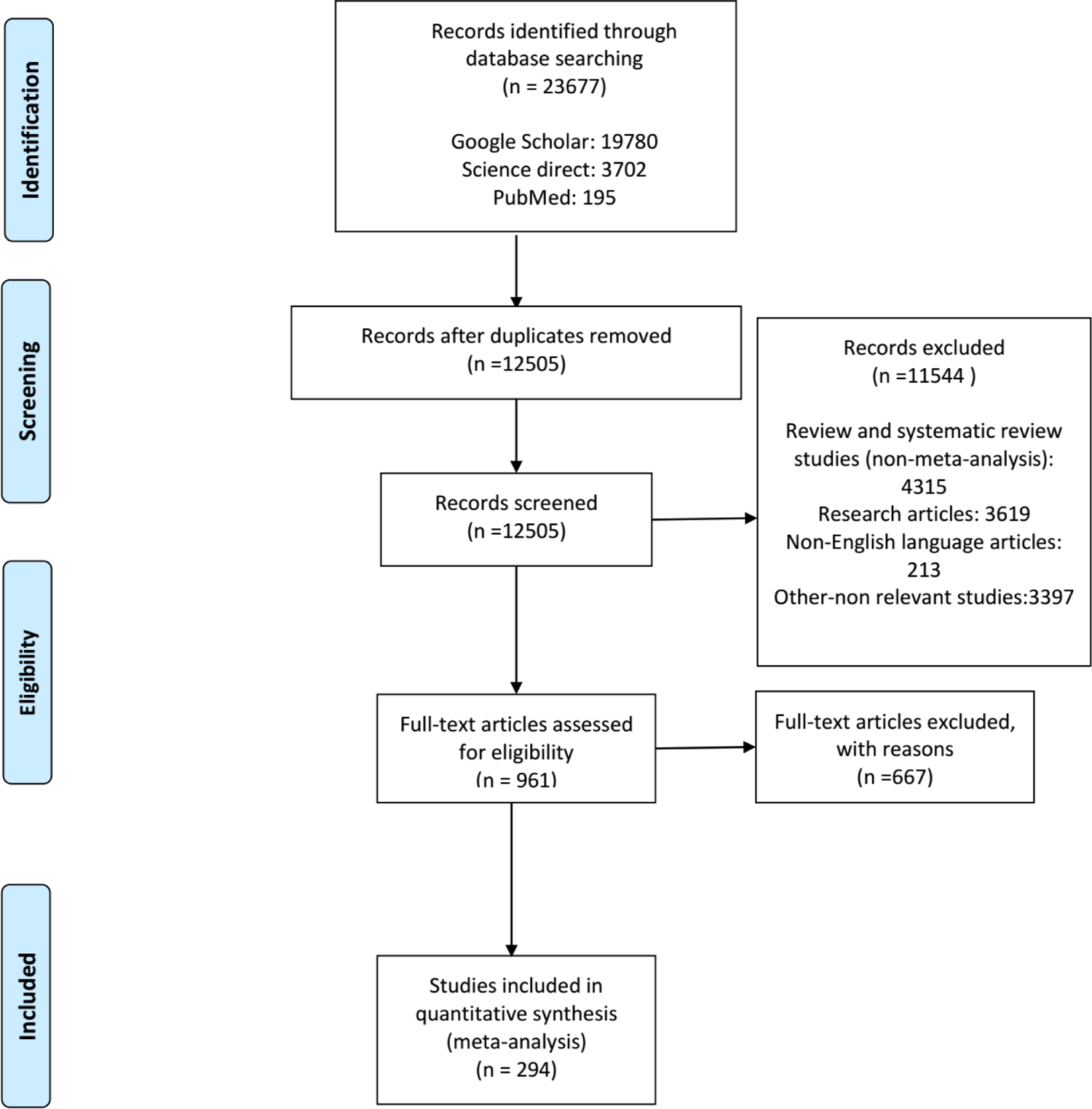
PRISMA flow diagram of search strategy and study selection.

### Distribution of articles by gender

In the distribution of articles that explicitly referred to men or women being tested, five articles were devoted to women (5–9), while no articles specifically focused on the role of probiotics on men’s diseases. The specific researches for the women are mainly based on their specific attributes like pregnancy or vaginal health such as bacterial vaginosis, and polycystic ovary syndrome(5–9).One study reported on urinary tract infection(5),although this manifests itself in both sexes, it is more prevalent in women.

### Distribution of articles by age

Figure 2 shows the distribution of articles by different age groups. Statistics are only based on the studies that clearly mentioned the age groups in their meta-analysis. As can be seen in the figure, most studies have been done on infants and children, and these two age groups alone account for 63% of the total studies.

**Figure 2.**
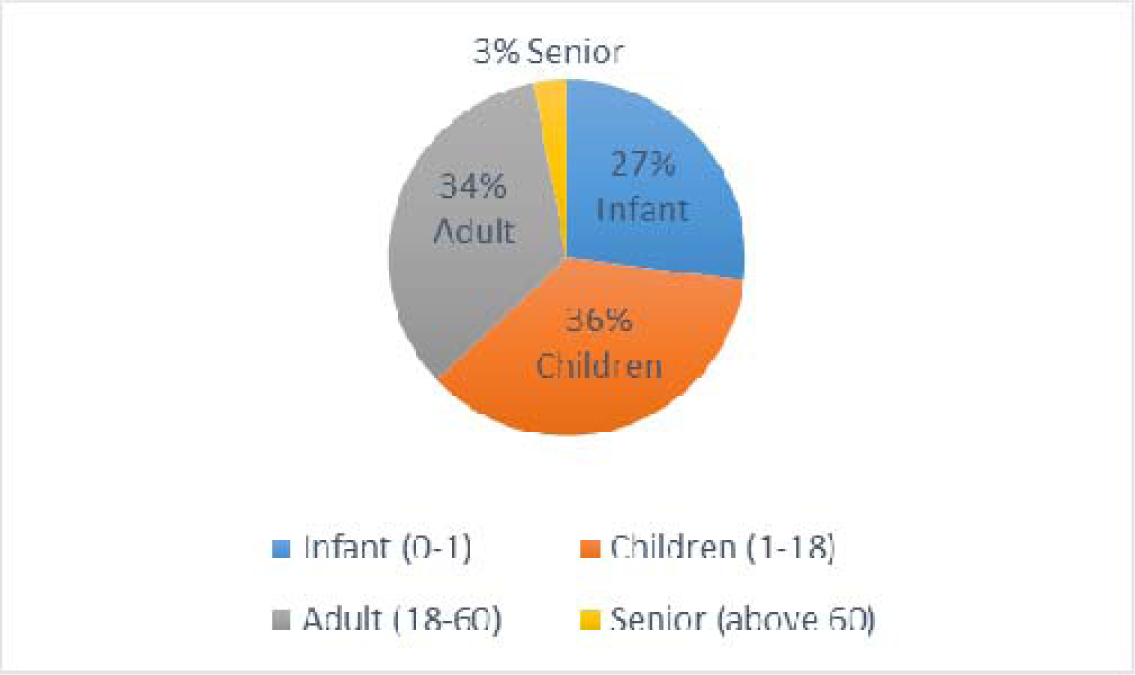
Distribution of probiotics-disease related meta-analysis articles by age category.

### Distribution of articles based on the effectiveness report

As shown in Figure 3, 79% of the meta-analysis studies showed an overall positive effect of probiotics. No analysis was found that reported a negative effect of probiotics in the prevention and treatment of various diseases, and only 21% of the included studies reported probiotics had no effect on the studies disease.

**Figure 3.**
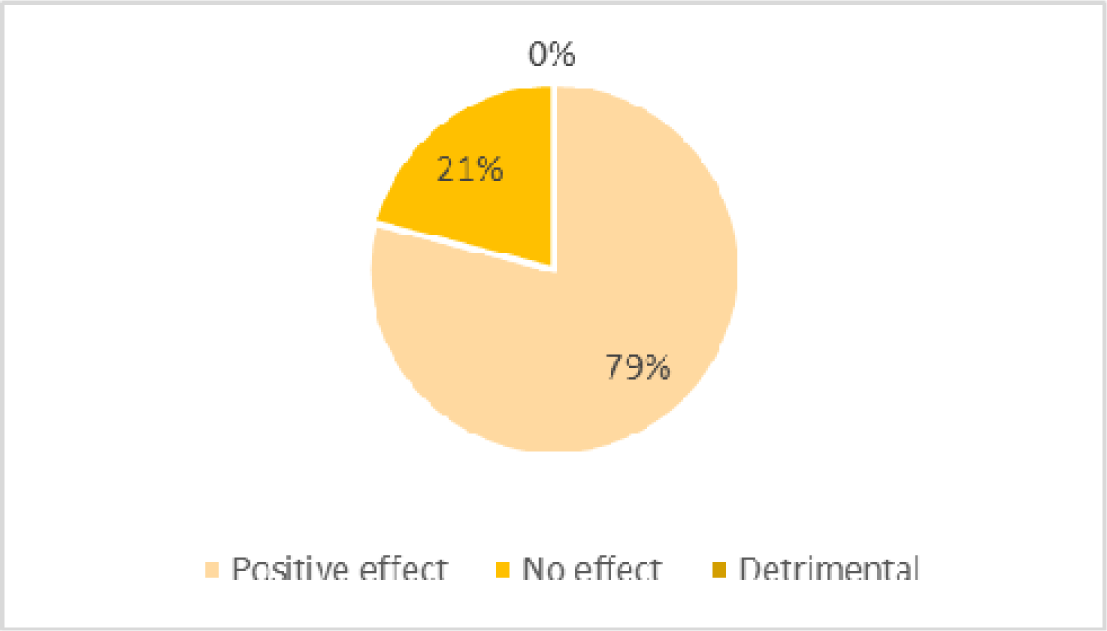
Meta-analyses distribution based on efficacy of the used probiotics.

In Figure 3 the percentage of probiotics effectiveness are shown by the most studied diseases. However, probiotics have had a 100 percent effect on a number of other diseases such as treatment of Colic, prevention and treatment of Atopic Dermatitis, prevention of Eczema, treatment of Cutaneous wounds, prevention of Common Cold, prevention of cancer, treatment complications including diarrhea, prevention of Atopy, treatment of Inflammation, treatment of Critical illness, treatment of Psychological Stress, treatment of Bacterial vaginosis, prevention of Urinary tract infections, treatment of Diabetes especially diabetes Type 2, prevention and treatment of Hepatic encephalopathy, remission induction and maintenance effect on Ulcerative colitis, prevention and treatment of gastrointestinal disorder, treatment of Pouchitis, prevention of Colorectal surgery, prevention and treatment of Lipid profile, treatment of Blood pressure, prevention of Intraventricular Hemorrhage, prevention of Sepsis, prevention of Oral candidiasis, treatment of Pathological Neonatal Jaundice, treatment of *Helicobacter pylori* infection(eradication therapy).

When defining “efficacy” as the effectiveness of probiotics on curing or improvement of the disease based on the result of each meta-analysis study, Figure 4 shown the success rate. Diarrhea in Figure 4 refers to infectious and non-infectious causes such as antibiotic-related diarrhea, travel-related diarrhea, acute diarrhea, radiation-induced diarrhea, *Clostridioides difficile*-Associated Diarrhea, cancer therapy-induced diarrhea, acute rotavirus diarrhea, etc. Diabetes refers predominantly to type 2 diabetes. However, some articles did not mention a specific type of diabetes and generally investigated the effect of probiotics on diabetes. Two cases also addressed gestational diabetes. Prematurity is classified as a condition, based on ICD-10. Most articles on premature infants have studied necrotizing enterocolitis, but other cases including sepsis, intraventricular hemorrhage, mortality, length of hospital stay, and weight gain. So, we decided to consider all of them as preterm infant. Infections also includes respiratory infections, *C. difficile* infections, and *Helicobacter pylori* infections.

**Figure 4.**
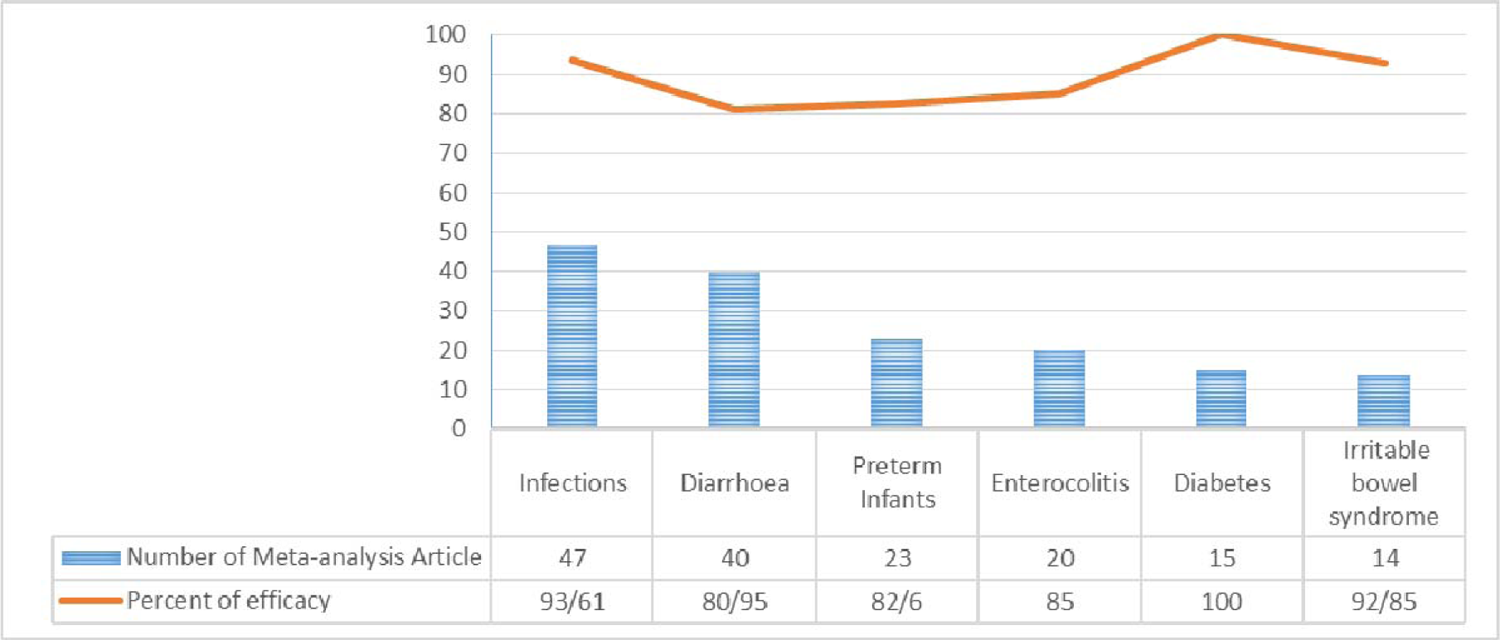
Efficacy percentage of probiotics on diseases with the highest number of studies.

Out of the 283 identified articles, 57 reported no effect of the probiotics in preventing or treating diseases, Table 1. Among them; Inflammatory Bowel Disease including Crohn’s Disease, Acute or special types of diarrhea and Asthma have the most cases. Actually, all of the Asthma related meta-analysis probiotic studies report ineffectiveness. Maybe one other probable reason is that some papers, 25 out of 57, are based literally only on a handful of studies, less than 10 studies. Conflicting outcomes will also be discussed later in Table 3. Although probiotics have had a beneficial or neutral effect in preventing and treating diseases in humans, no negative effects of probiotics on human health have been found in this review of meta-analyses (10). Nevertheless, single studies may have observed rare adverse events, e.g. (11).

**Table 1.**
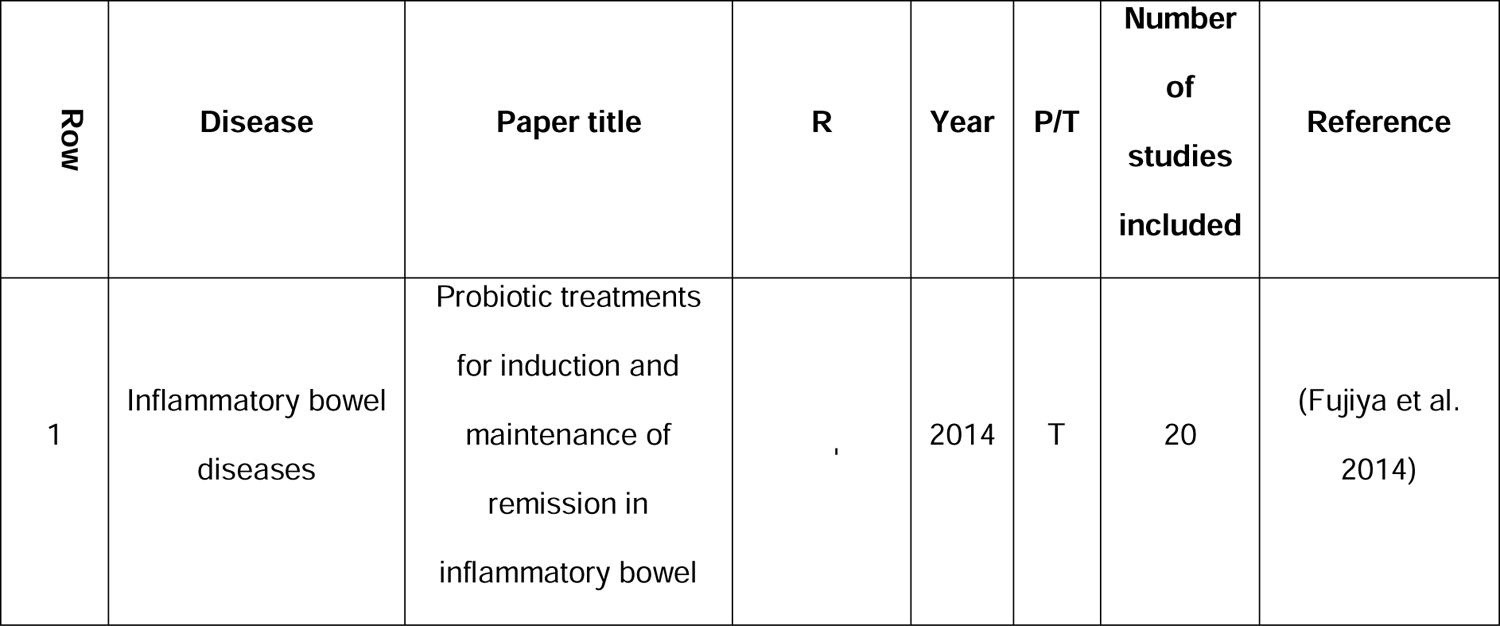
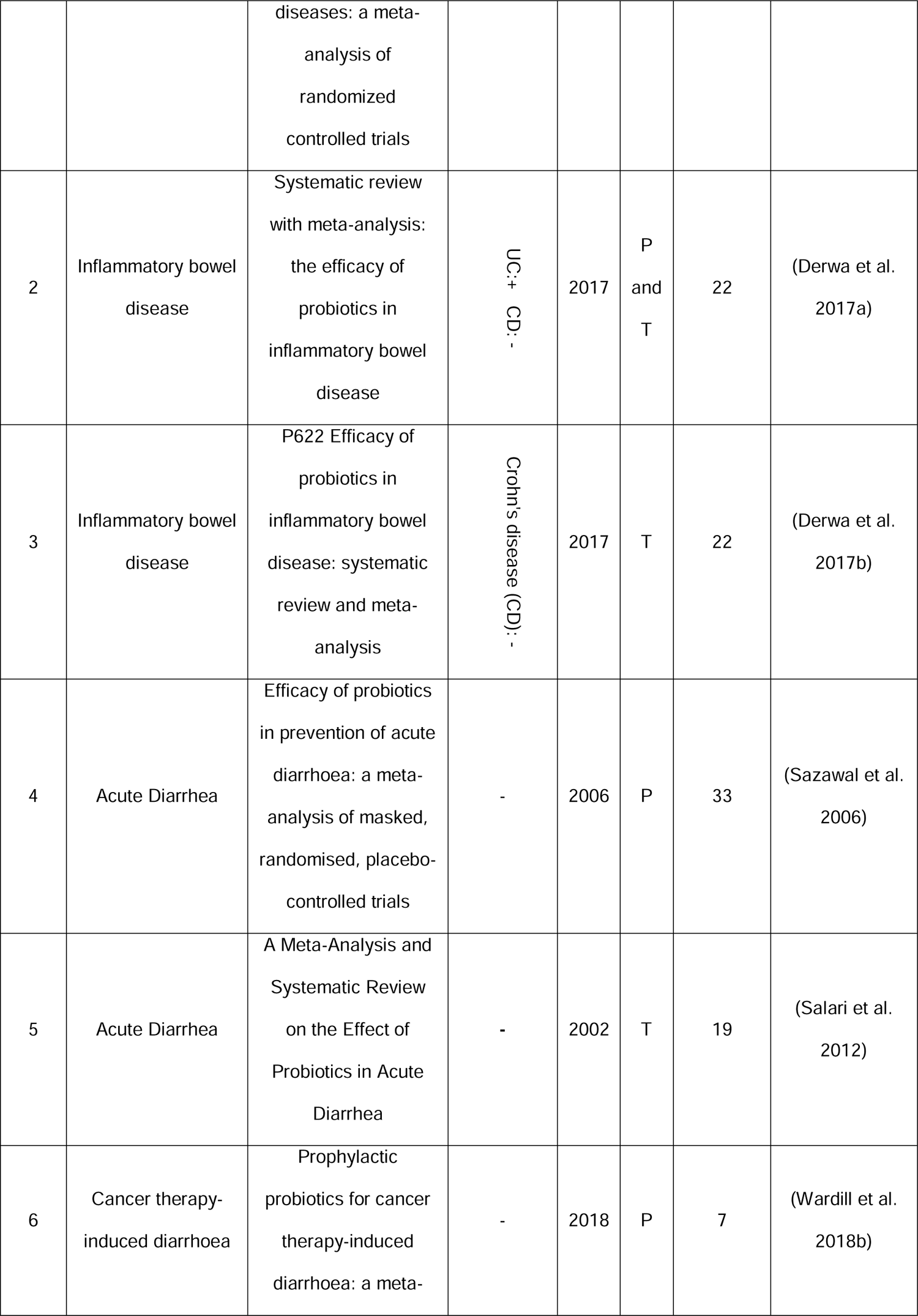
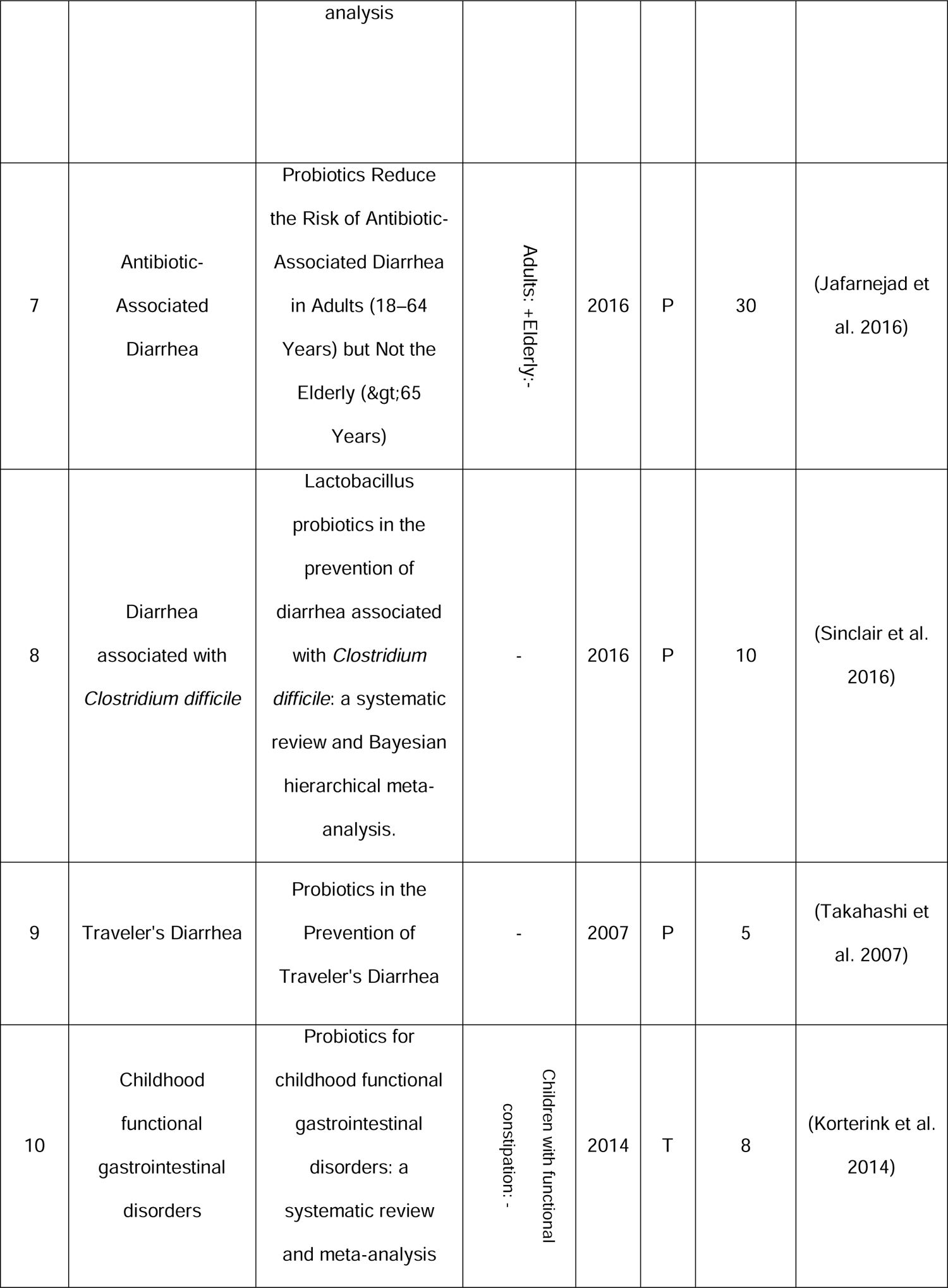
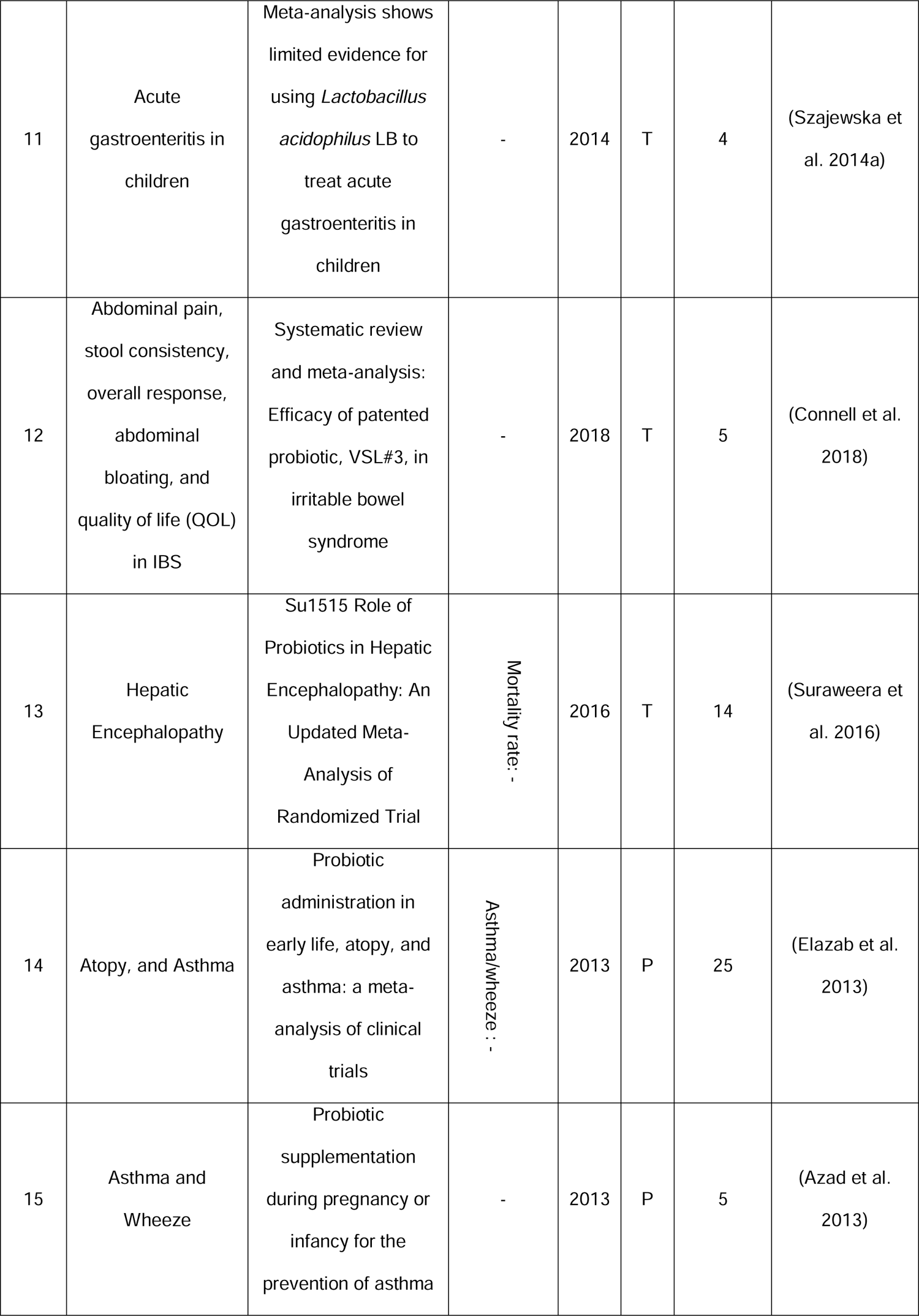
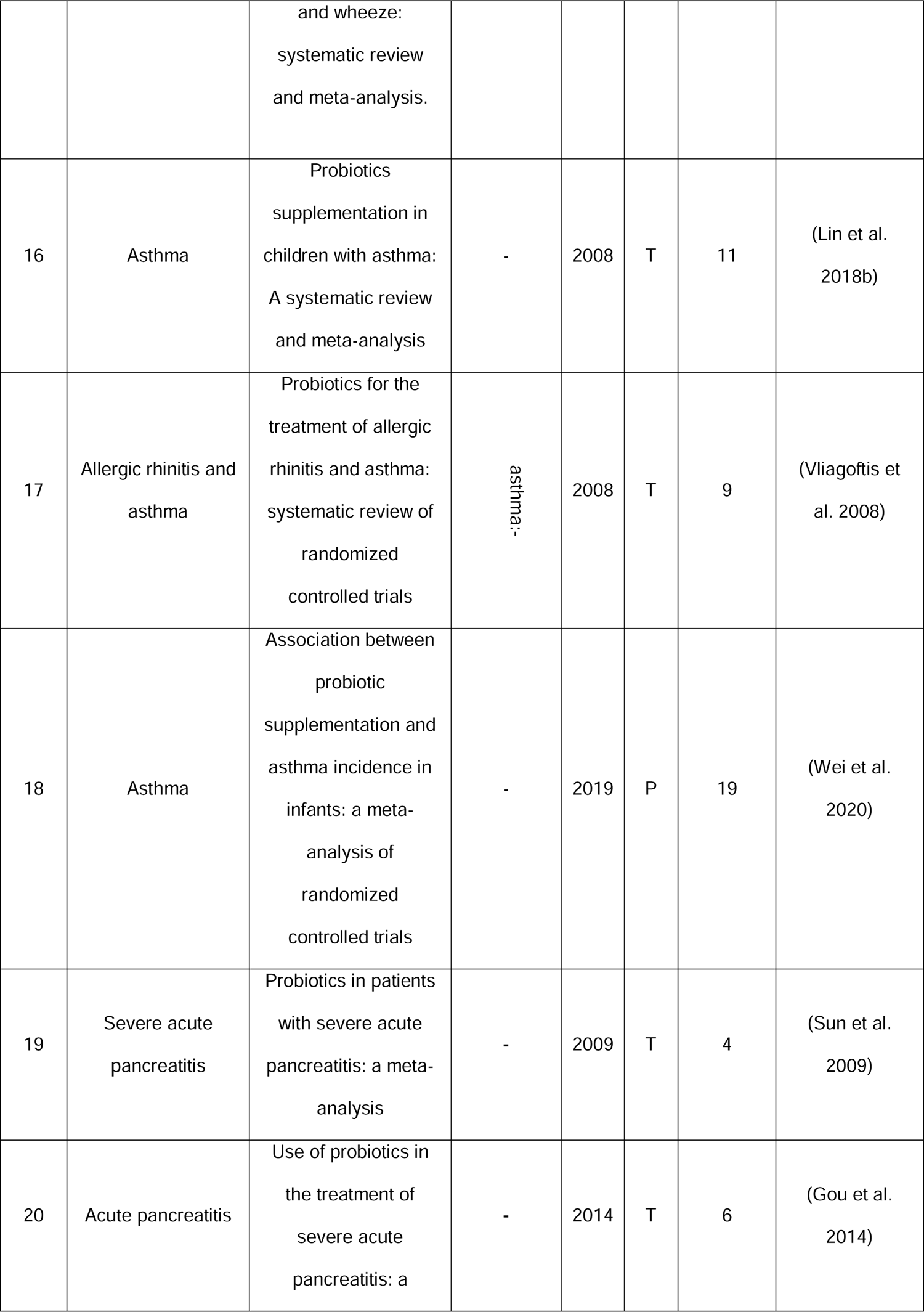
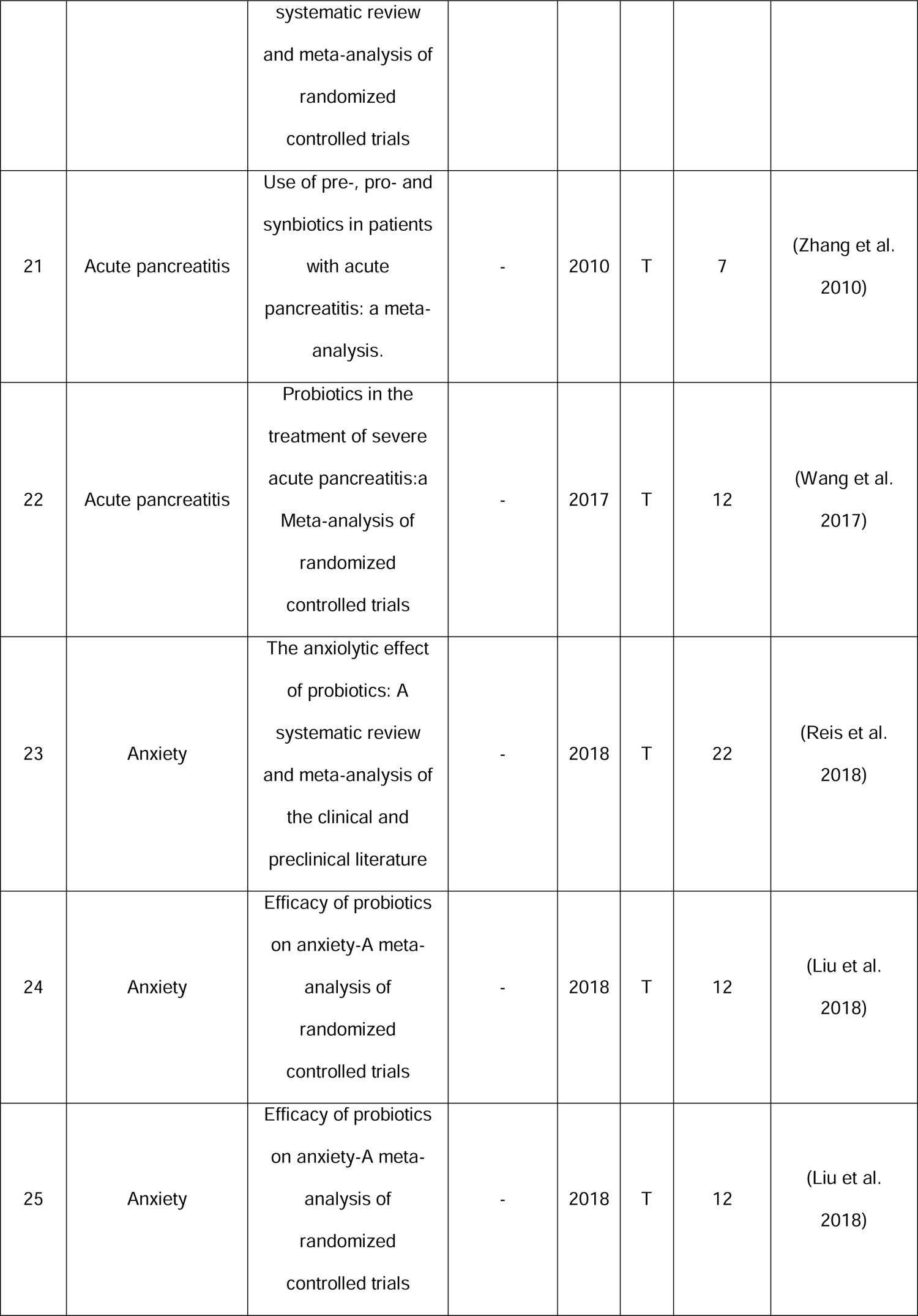
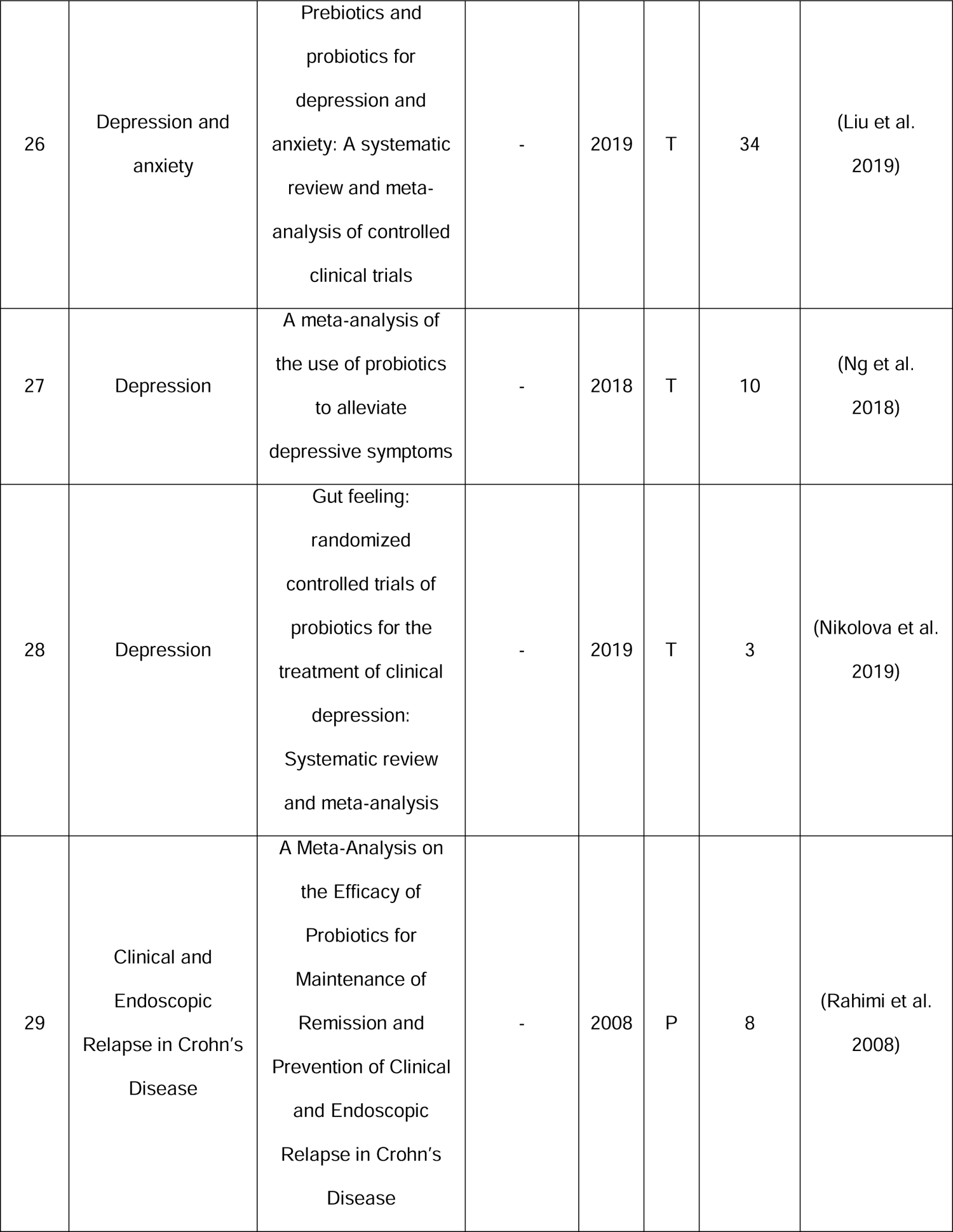
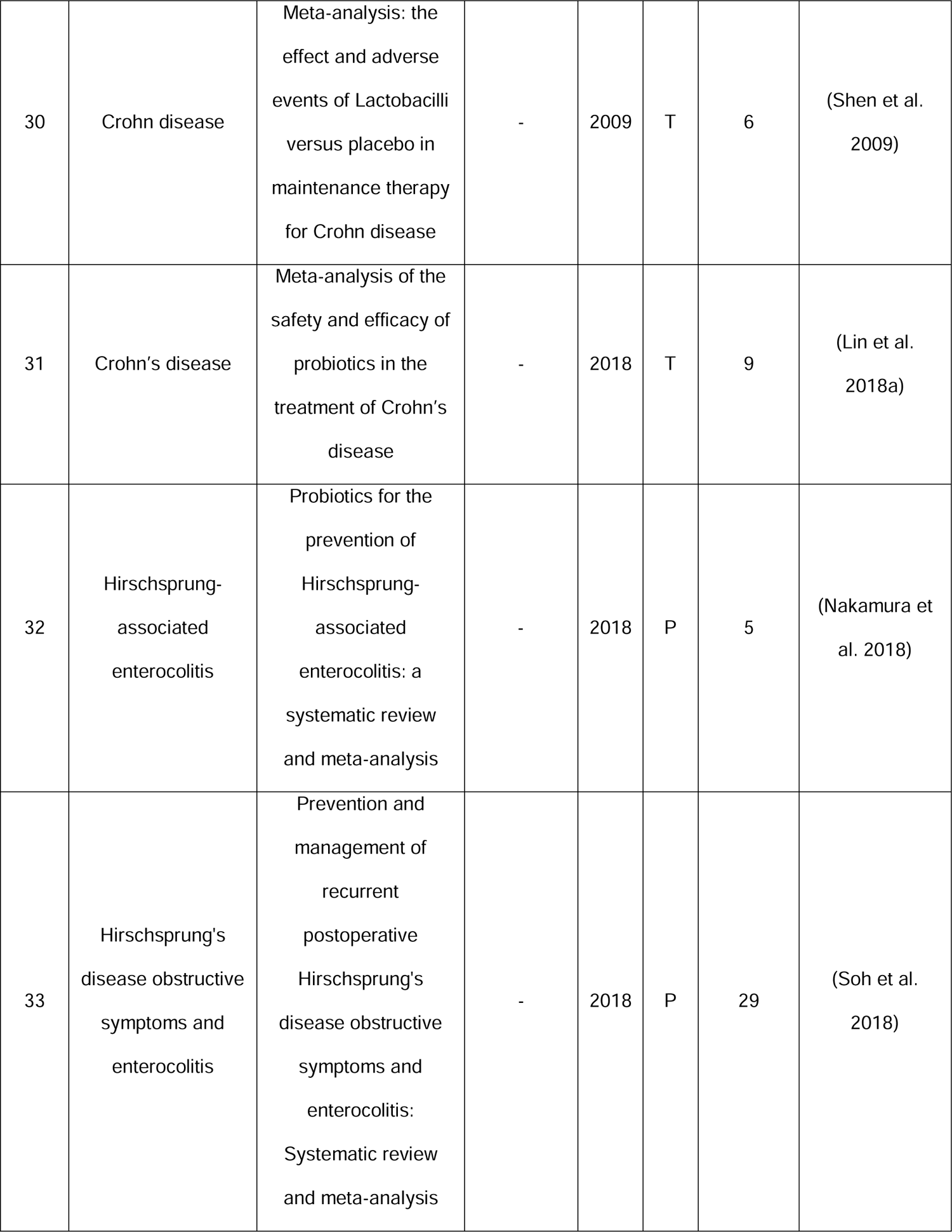
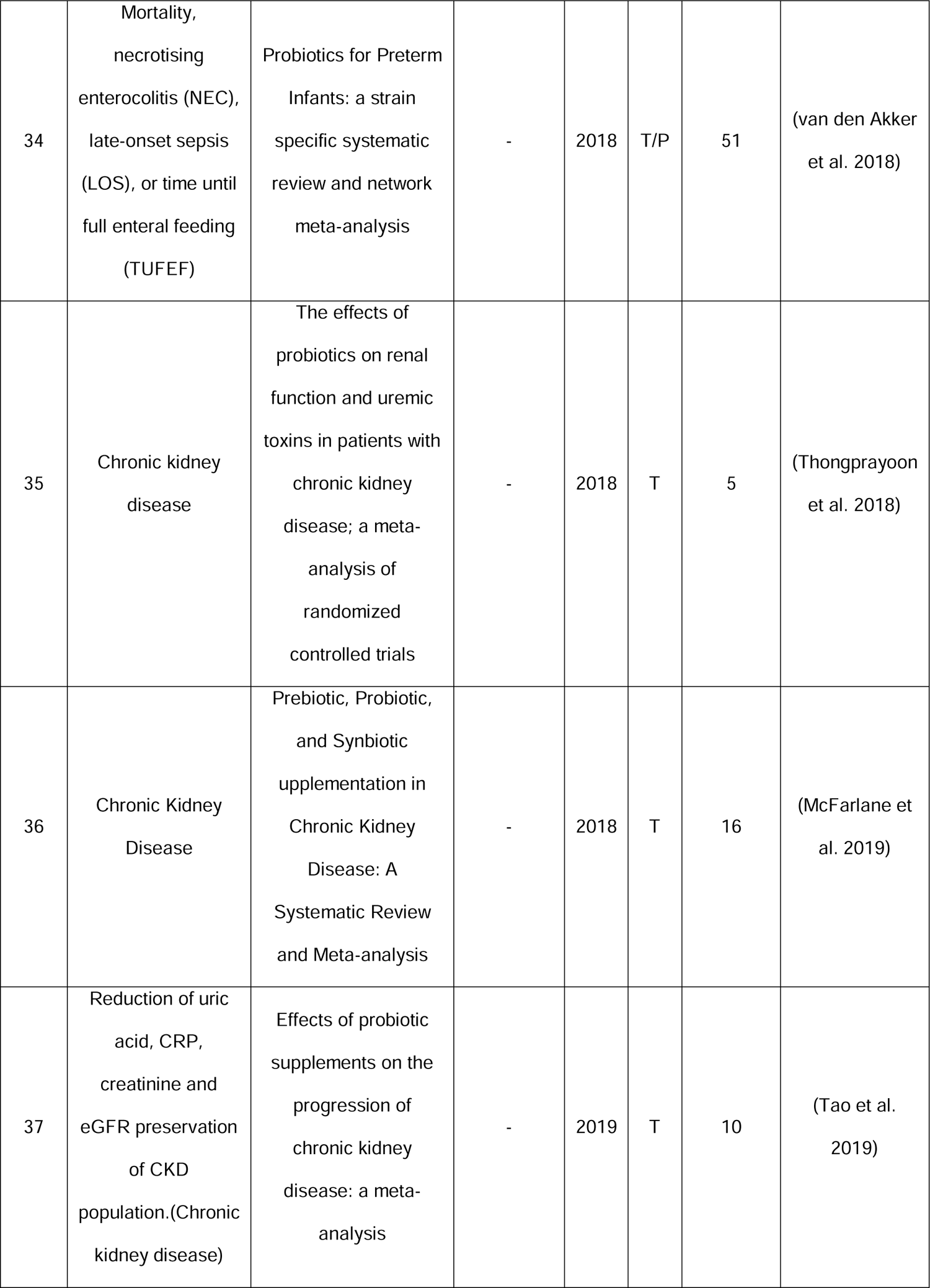
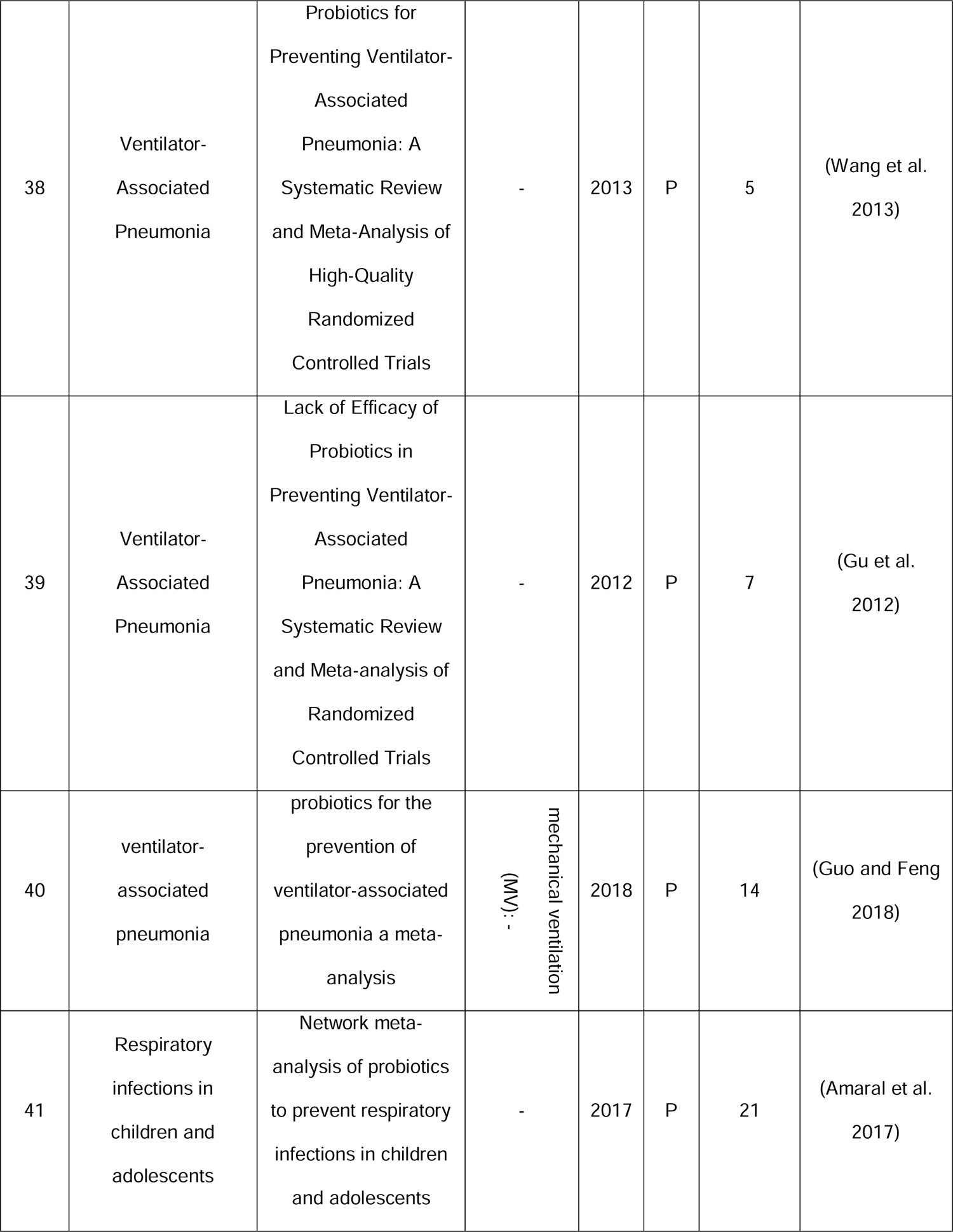
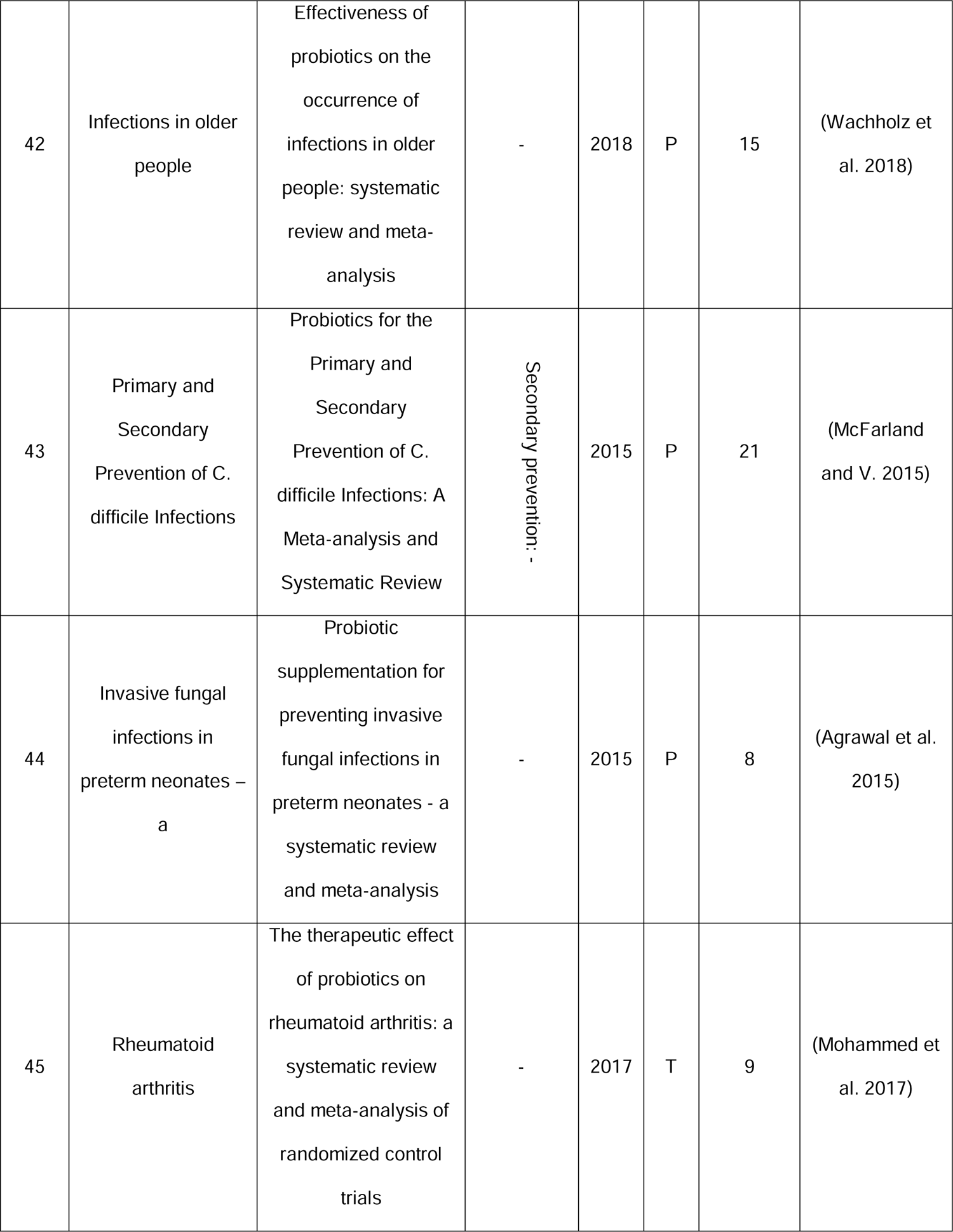
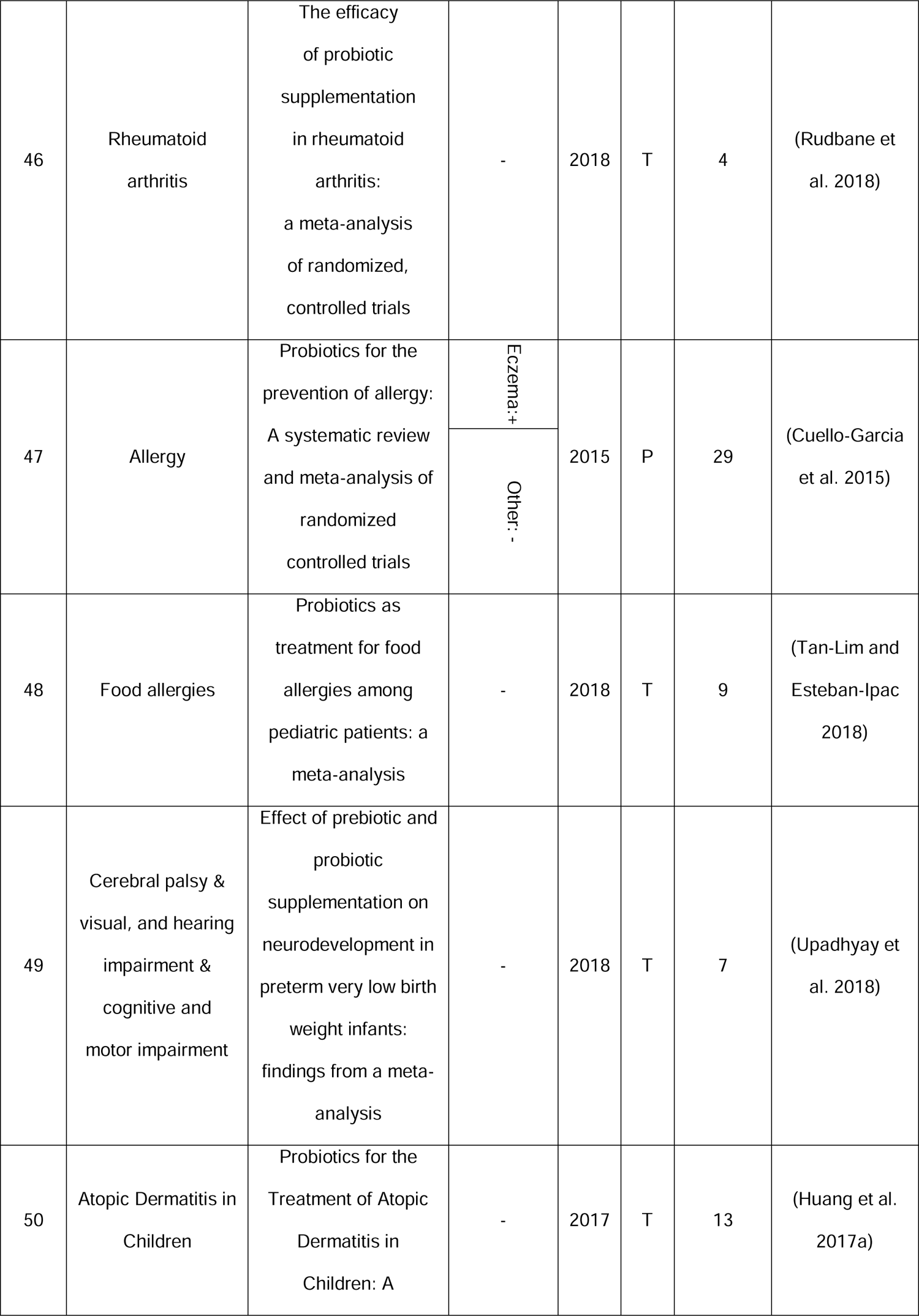
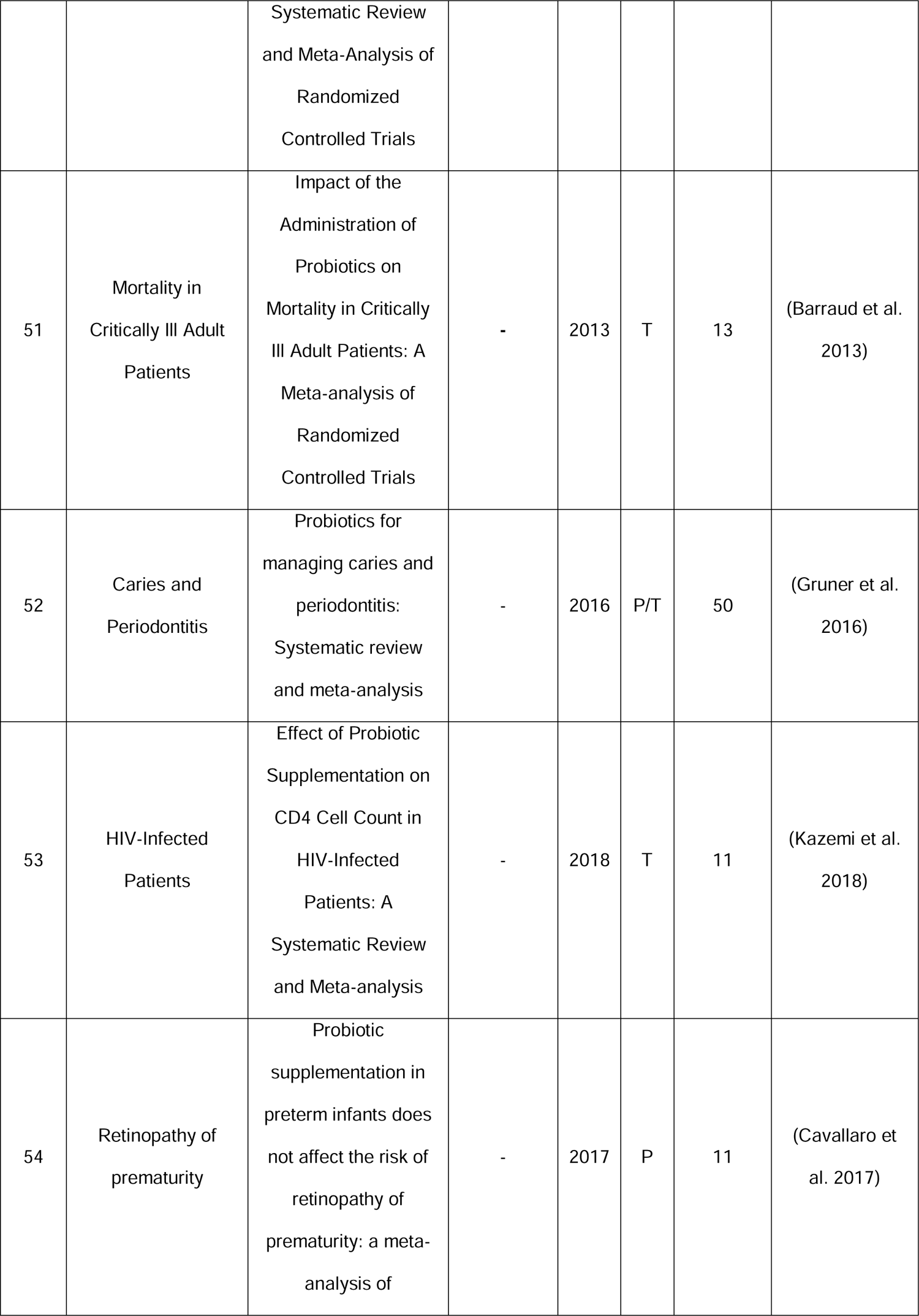
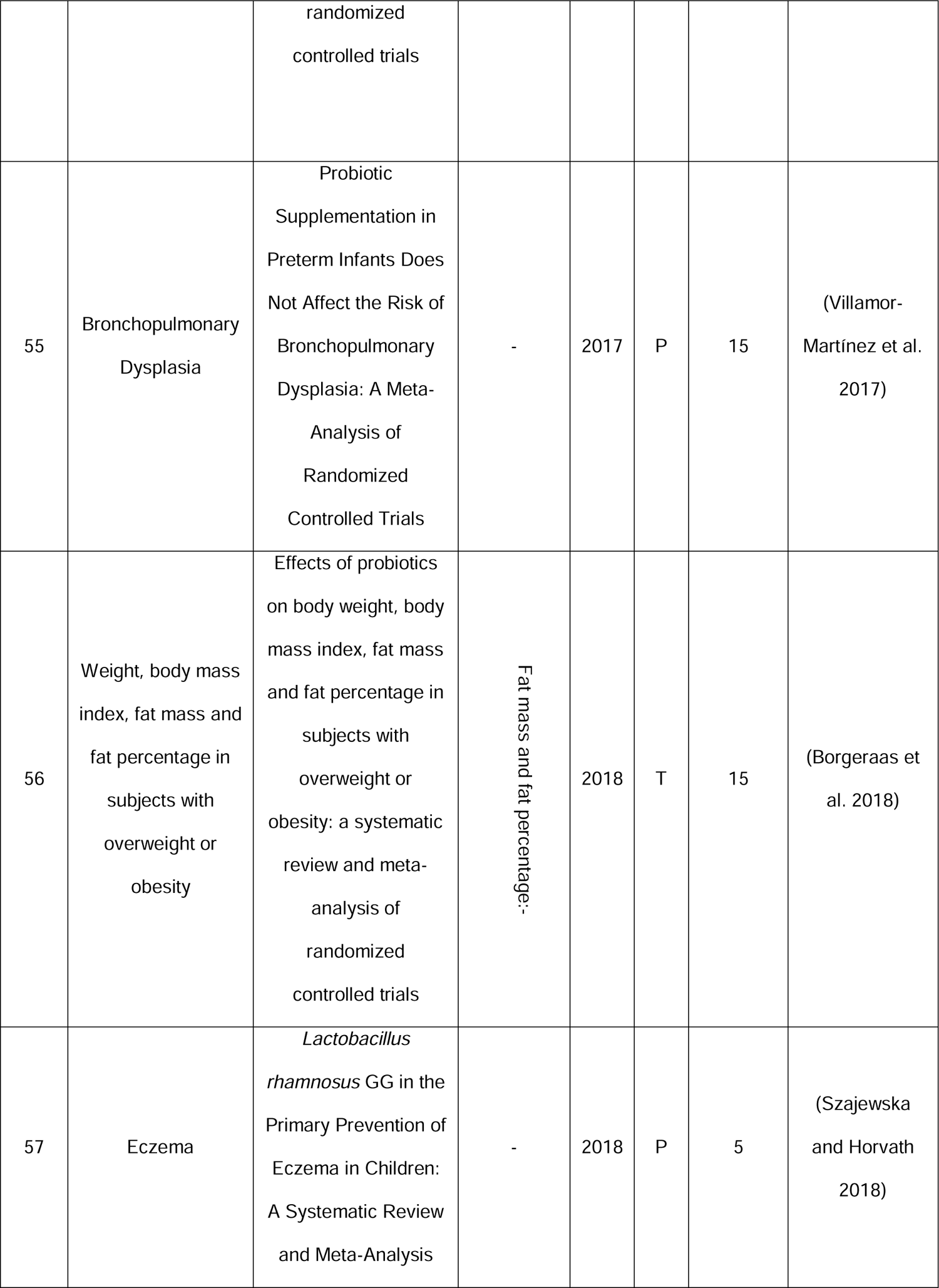
Ineffectiveness reports of probiotics for various diseases. R stands for Result and P/T is the abbreviation for Prevention/Treatment. Sign + indicates the **positive** effect of probiotics on the disease. Sign - Indicates that probiotics do not have a positive effect on patients (nor having a negative effect on patients).

### Distribution of articles by disease type

Figure 5 shows the distribution of articles by anatomical of physiological target. The highest number of articles were related to digestive system diseases (139 out of 283 or 49.11% of the total) and the least published articles were related to diseases of the nervous system, eye and adnexa. Percentage of the effectiveness of the probiotics in **treatment** of the disease is also shown in the figure. It is more than 50% for all the diseases. Apparently, the number of meta-analysis articles in this article is expected to be the same as the number of diseases, but the number of diseases reviewed is greater than the total number of articles. The reason is that some articles have reviewed several diseases. As a result, the number of diseases has exceeded the total number of articles.

**Figure 5.**
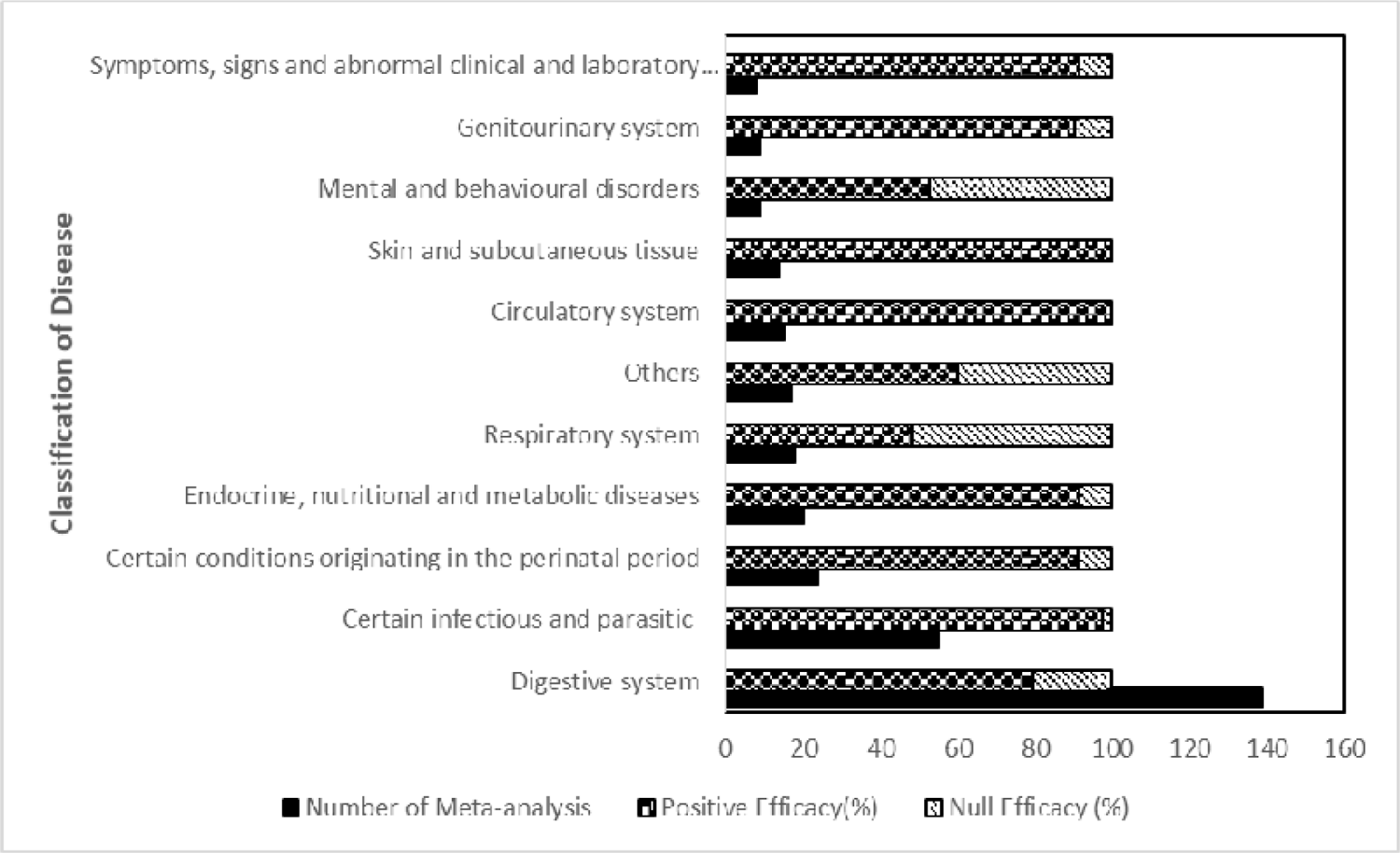
Distribution of probiotics-disease related meta-analysis articles based on the classification of diseases. The effectiveness of probiotics was defined as the percent overall curing or improvement of the disease based on the result of each meta-analysis study.

Figures 6 to 17 provide more detailed statistics for each disease category identified in Figure. 5. Extensive studies (139 cases) have been performed on the effect of probiotics on gastrointestinal diseases that encompass various gastrointestinal diseases (Figure 6). Interestingly, among of 20 studies on enterocolitis, 19 were related to necrotizing enterocolitis.

**Figure 6.**
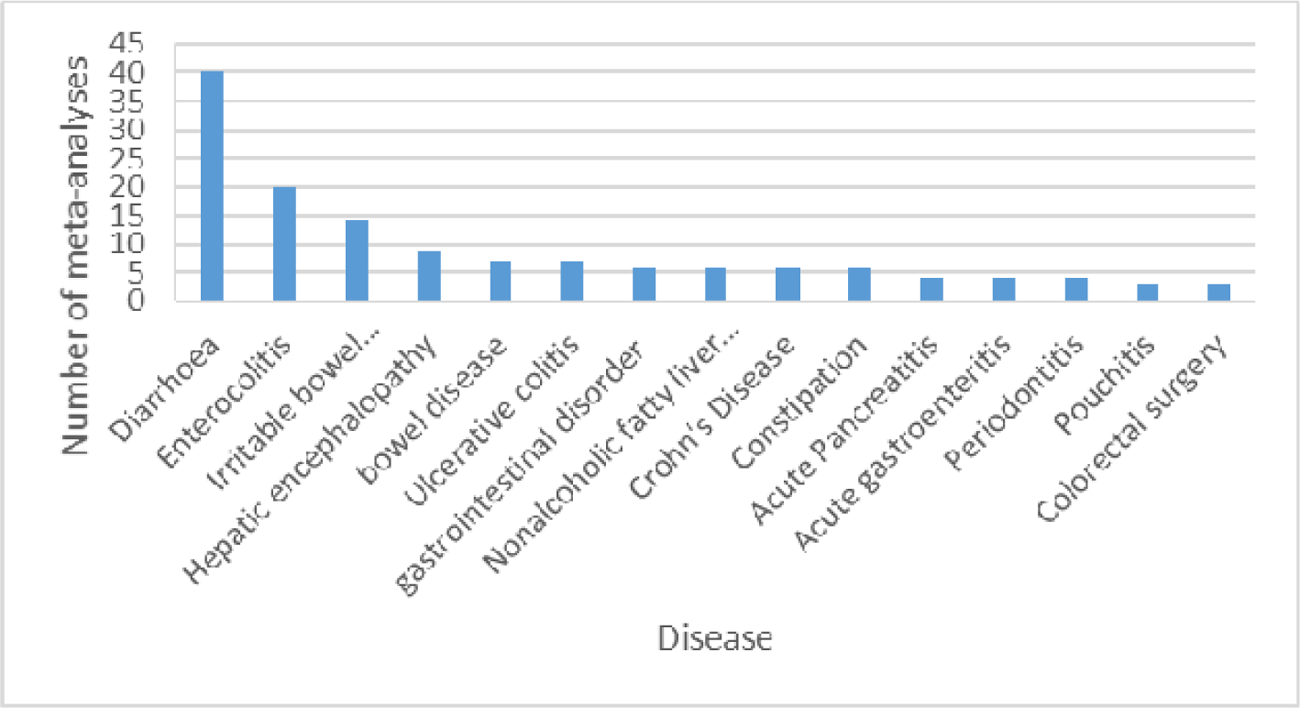
Distribution of meta-analyses based on Digestive system’s diseases.

Most of studies included in this review concern digestive system’s diseases, and among the total number of them, the diarrhea has the highest number of analyses (40 articles). Several types of diarrhea that have been studied in these meta-analyses are including, antibiotic-related diarrhea, travel-related diarrhea, acute diarrhea, radiation-induced diarrhea, *C. difficile*-Associated Diarrhea, cancer therapy-induced diarrhea and acute rotavirus diarrhea.

Probiotics have a beneficial role in intestinal function. They enhance tight intestinal connections and enhance intestinal mucosal barrier’s action by increasing mucus production. They also reduce inflammation and return the normal bowel movements. As a result, probiotics with these mechanisms play an important role in the management of gastrointestinal diseases (12, 13). According to our results, probiotics have been able to be quite effective in the prevention and treatment of diseases including irritable bowel syndrome, ulcerative colitis, pouchitis, necrotizing enterocolitis and constipation, and this finding can verify our assumption that probiotics can have a positive effect on the treatment or prevention of diseases especially for most of the gastrointestinal diseases.

Of the studies on infections (47 cases), 18 were *H. pylori* infection, 6 were respiratory infections and, 7 were *C. difficile* infections (figure 7). Based on the results, the most frequent meta-analysis studies were on diarrhea (40 articles, 14.13% of total studies). The results show that probiotics play an important role in the prevention and treatment of diarrhea, with 80.95% of the articles confirming its positive effect. Reports indicate that two probiotic strains that can be very effective in treating acute diarrhea in children are *Limosilactobacillus reuteri* ATCC 55730 (14, 15) and *Saccharomyces boulardii* (16, 17).

**Figure 7.**
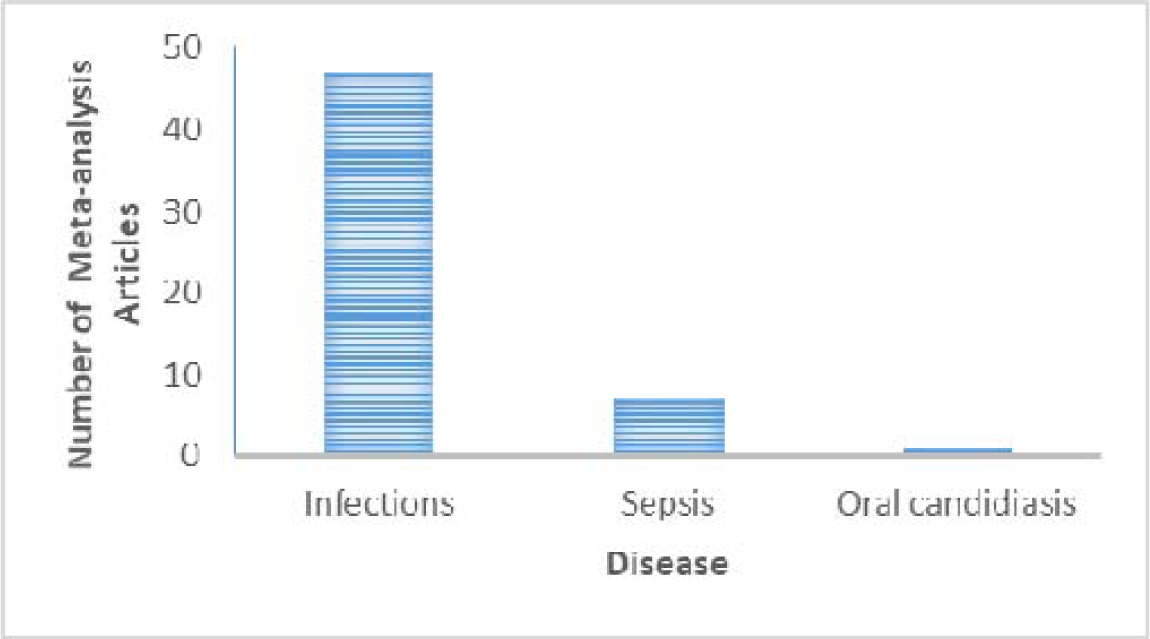
Distribution of meta-analysis articles based on disease of certain infectious and parasitic.

In fact, probiotics work through a variety of mechanisms, including directly eliminating or preventing pathogens growth, destroying toxins, competing against target cells, or regulating the immune system in these patients, and they also reintroduce the microbiota balance (the most common cause of different types of diarrhea is the disruption of this balance). Probiotics have a prophylactic or therapeutic effect in all types of diarrhea, and its effectiveness is directly related to the type of strain, the antimicrobial and anti-inflammatory properties of the strain and its dosage (18–20).

Jaundice and the birth of a premature infant are rooted in pregnancy. After diarrhea, most of the meta-analysis articles have been published about premature infants and related diseases (Figure 8). Probiotic supplements administration is one of the most important therapeutic interventions in premature infants (21–30). Probiotics can absorb natural flora in these infants (31). In addition, it is reported that using probiotics may help preventing preterm labor also (32). According to the results, probiotics were 78.26% effective in preventing and treating various diseases in premature infants.

**Figure 8.**
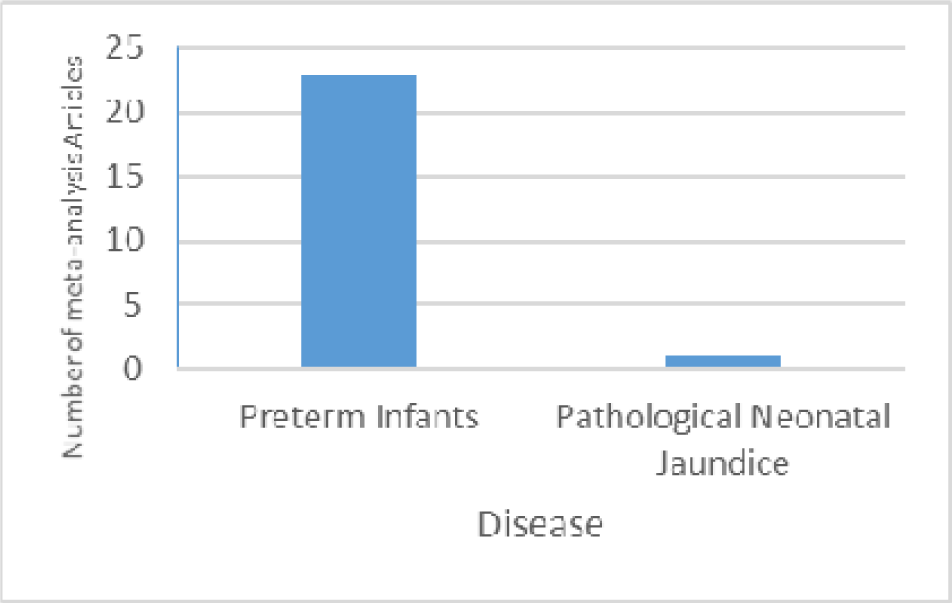
Distribution of meta-analysis articles based on certain conditions originating in the perinatal period.

The effect of probiotics on respiratory tract diseases is shown in Figure 9. Most of the respiratory diseases studied are pneumonia. Out of 6 studies, 5 were related to ventilator-associated pneumonia and 1 study was to nosocomial pneumonia.

**Figure 9.**
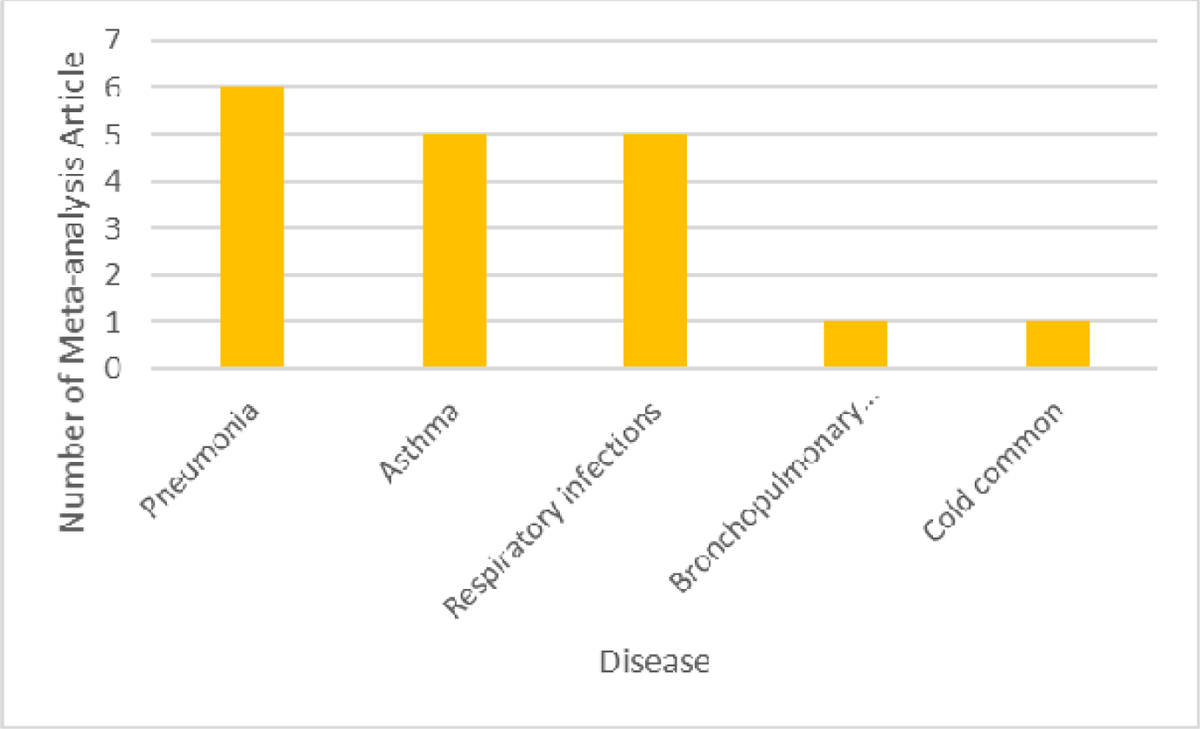
Distribution of meta-analysis articles based on Respiratory system’s disease.

According to the results, the percentage of probiotics effectiveness in respiratory infections is 83.33%. One of the reasons that probiotics reduce the risk of infectious diseases, including respiratory infections, is the functional role of these strains in the gastrointestinal system as well as their association with the immune system (33), Which stimulate the immune system and strengthen it (34, 35). Probiotics have also been 66.66% effective in preventing ventilator-associated pneumonia, which is an acute respiratory infection and the highest cause of death in children worldwide (36).

Within metabolic and endocrine diseases, 9 out of 15 articles were related to type 2 diabetes, Figure 10. Probiotics have been quite effective in controlling diabetes, especially type 2 diabetes. In diabetic patients, antioxidant defenses become disturbed and large amounts of free radicals are produced (35, 37), and since oxidative stress plays an important role in the pathogenesis, progression, or complications of diabetes (35,38,39), probiotics by their antioxidant properties, can reduce inflammation and also prevent the destruction of pancreatic beta cells, thereby reducing blood sugar (40, 41). Because most meta-analysis studies have been done on type 2 diabetic patients, more studies are recommended to investigate the effect of probiotics on type 2 diabetic patients.

**Figure 10.**
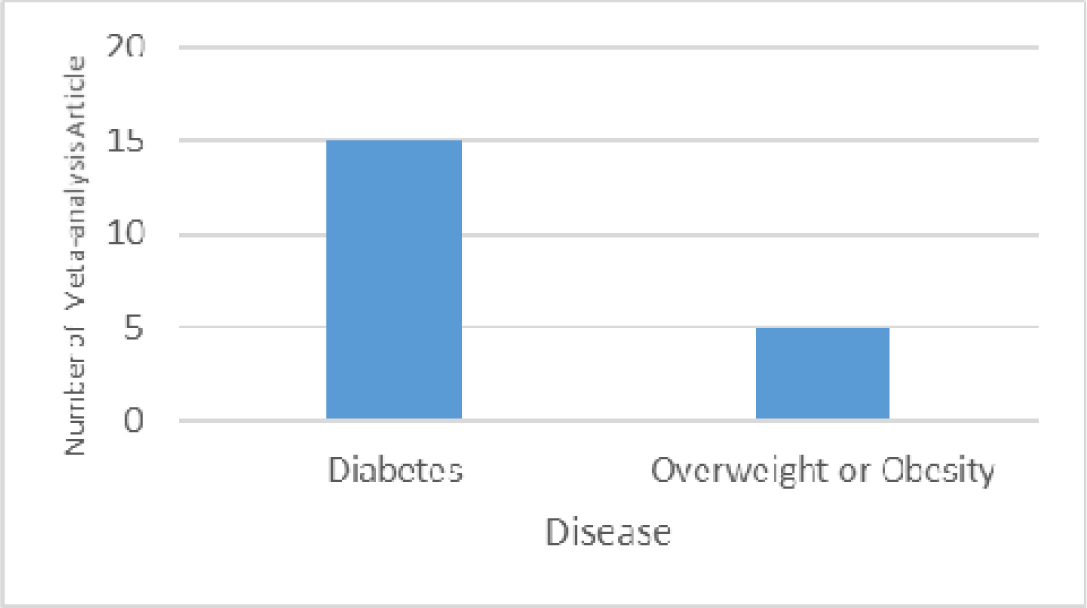
Distribution of meta-analysis articles based on Endocrine, nutritional and metabolic’s diseases.

Only five meta-analysis articles have examined the effect of probiotics on obesity and overweight (42–46). However, in all cases, except one (42), probiotics had a positive effect on weight and body mass index (BMI) improvement. Other meta-analysis studies have investigated the effect of probiotics on body weight and BMI, which reported different results. Therefore, further studies are needed to determine the precise effect of probiotics on human weight. A study by Zhang and colleagues suggests that probiotics, especially when used multiple probiotic strains in combination for more than 8 weeks, can reduce weight and also decrease body mass index (47). Another study reported that probiotics had no effect on weight loss(48). Million and colleagues have also reported that different species of *Lactobacillus* have different effects on weight change (49).

Intestinal microbiota is one of the contributing factors in obesity (50, 51) an increase in the strains of *Bacteroides fragilis*, *Clostridium leptum*, and *Bifidobacterium catenulatum* and decrease in the levels of *Clostridium coccoides*, *Lactobacillus sensu lato*, and *Bifidobacterium* have significant effective on weight loss (52, 53) so probiotics may be effective in modulating obesity by altering the composition of the gut microbiota (54, 55).

Probiotics that are effective in controlling obesity and appetite exert their effects through the nervous system. Lactobacilli and bifidobacteria, which are common probiotics, are capable of producing short chain fatty acids (SCFAs) (56, 57). They have positive effects on energy metabolism in mammals (58) which decrease appetite by altering the levels of glutamate, glutamine, gamma amino butyric acid (GABA) and neuropeptides. On the other hand, acetate, the short-chain fatty acid produced by bifidobacteria, causes physiological and regulatory changes in the hypothalamus resulting in suppression of appetite (59–61)

There have also been 14 studies on the effects of probiotics on skin diseases that are illustrated in Figure 11 for their distribution in different types of skin diseases. According to the results of this study, probiotics have been very effective in preventing eczema. However, Doege and colleagues have shown that probiotic are effective in preventing these diseases only when *lactobacilli* strains are used during pregnancy, not a combination of different probiotic strains (62).

**Figure 11.**
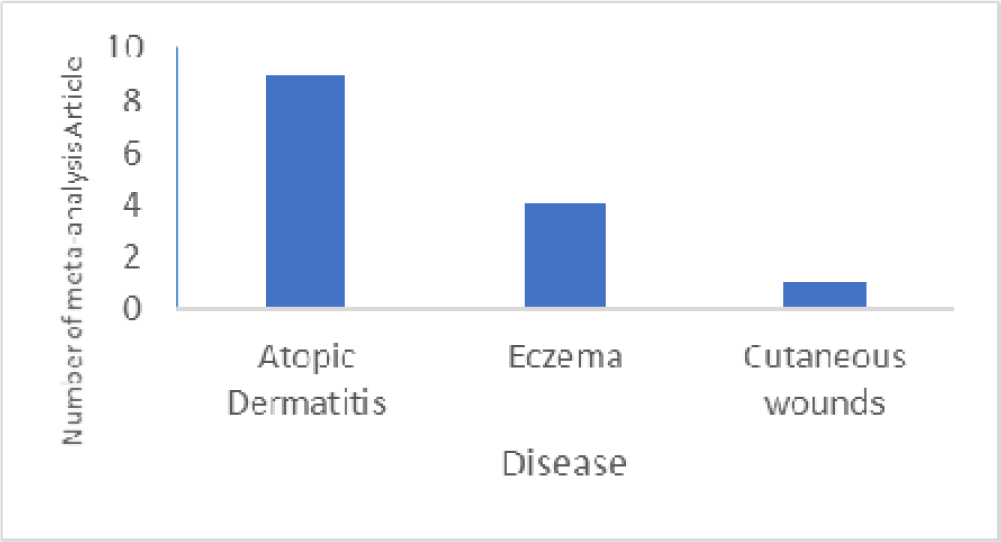
Distribution of meta-analysis articles based on Skin’s disease.

According to the International **Classification** of Diseases, mortality and colic are classified as symptoms, signs and abnormal clinical and laboratory findings. The number of articles published in this field so far is shown in Figure 12.

**Figure 12.**
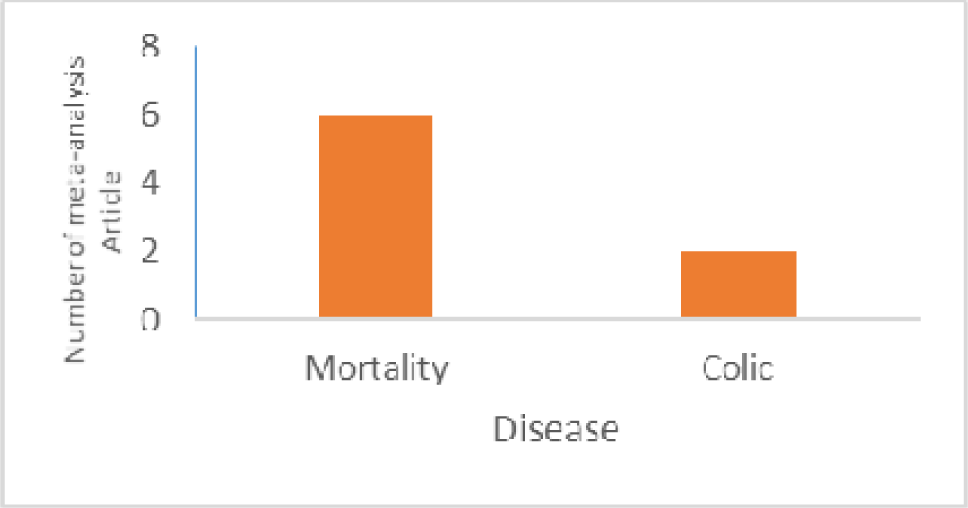
Distribution of meta-analysis articles based on symptoms, signs and abnormal clinical and laboratory findings.

Figure 13 shows the circulatory system related diseases. The low number of studies in this area is clearly evident. According to the results presented in Table 2, probiotics also have a positive effect on reducing fat and **cholesterol**. In a meta-analysis study investigating the effect of probiotics on lipid profile and blood pressure in patients with type 2 diabetes, it was reported that probiotics may be a new method for lipid profile and management and control of blood pressure in patients with type 2 diabetes(63).

**Figure 13.**
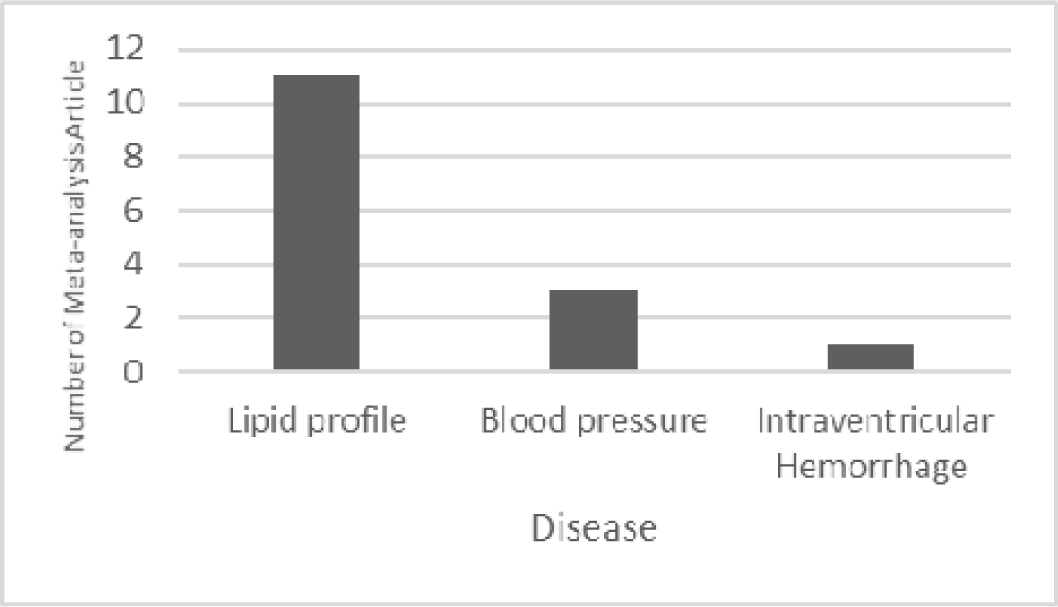
Distributed of meta-analysis articles based on Circulatory system’s diseases.

**Table 2.** Classification of the meta-analysis articles impact on human health apart from disease. R is the abbreviation of Result and P/T stands for Prevention/Treatment. Sign + indicates the positive effect of probiotics on the disease. Sign - Indicates that probiotics do not have a positive effect on patients (nor having a negative effect on patients) Table 3 presents articles that have studied a particular disease but reported different results. This contradiction may be due to the publication of articles in different years and the completion of scientific findings from new experiments and studies.

| Row | Case | Paper title | R | Year | P / T | The number of studies included | Reference |
| --- | --- | --- | --- | --- | --- | --- | --- |
| 1 | Oxidative stress | A systematic review and meta-analysis of the probiotics and synbiotics effects on oxidative stress | + | 2018 | T | 27 | (102) |
| 2 | Blood glucose | Effect of probiotics and synbiotics on blood glucose: a systematic review and meta-analysis of controlled trials | + | 2018 | T | 14 | (103) |
| 3 | Body weight and body-mass index | Effect of probiotics on body weight and body-mass index: a systematic review and meta-analysis of randomized, controlled trials | + | 2016 | T | 20 | (47) |
| 4 | Safety in pregnancy | Probiotic safety in pregnancy: a systematic review and meta-analysis of randomized controlled trials of Lactobacillus, Bifidobacterium, and Saccharomyces spp. | - for Lactobacillus and Bifidobacterium.<br>The safety of Saccharomyces is unknown | 2009 | T | 11 | (104) |
| 5 | Outcomes of Trauma | The Effects of Probiotics in Early Enteral Nutrition on the Outcomes of Trauma | Nosocomial infections, VAP, and length of ICU stay: +<br>Mortality: - | 2013 | T | 5 | (105) |
| 6 | Weight gain in humans and animals | Comparative meta-analysis of the effect of Lactobacillus species on weight gain in humans and animals | Different Lactobacillus species are associated different effects on weight change | 2012 | T | 82 | (49) |
| 7 | Metabolic | The effects of | Probiotics | 2018 | T | 10 | (106) |
|  | health in pregnant women | probiotics supplementation on metabolic health in pregnant women: An evidence based meta-analysis | supplementation have beneficial effects on glucose metabolism, rather than lipid metabolism |  |  |  |  |
| 8 | Immune response to influenza vaccination in adults | Effect of probiotics and prebiotics on immune response to influenza vaccination in adults: A systematic review and meta-analysis of randomized controlled trials | + | 2018 | T | 9 | (107) |
| 9 | Antibody titers after influenza vaccination | The influence of prebiotic or probiotic supplementation on antibody titers after influenza vaccination: a systematic review and meta-analysis of randomized controlled trials. | + | 2018 | T | 12 | (108) |
| 10 | Weight loss | Probiotics for weight loss: a systematic review and meta-analysis | - | 2015 | T | 4 | (48) |
| 11 | Patients with severe head injury | Early enteral nutrition supplemented with probiotics improved the clinical outcomes in patients with severe head injury: protocol for a meta-analysis of randomized controlled trials | + | 2017 | T | 4 | (109) |
| 12 | Halitosis | The Effect of Probiotics on Halitosis: a Systematic Review and Meta-analysis | + | 2017 | T | 3 | (110) |
| 13 | Cellular immune function | Short-term probiotic supplementation enhances cellular immune function in healthy elderly: systematic review and meta-analysis of controlled studies | + | 2019 | T | 17 | (111) |
| 14 | Oral health | Are dairy products containing probiotics beneficial for oral health? A systematic review and meta-analysis | + | 2018 |  | 32 | (112) |
| 15 | Reduced antibiotic use | Does probiotic consumption reduce antibiotic utilization for common acute infections? A systematic review and meta-analysis | + | 2019 | T | 17 | (113) |
| 16 | Supplementat<br>iof Glucose<br>Metabolism<br>and Lipid<br>Profiles | The Effects of<br>Synbiotic<br>Supplementation on<br>Glucose Metabolism<br>and Lipid Profiles in<br>Patients with<br>Diabetes: a<br>Systematic Review<br>and Meta-Analysis of<br>Randomized<br>Controlled Trials | + | 2018 | T | 7 | (114) |
| 17 | Prevalence of<br>obesity | Effects of probiotic<br>supplementation on<br>the regulation of<br>blood lipid levels in<br>overweight or obese<br>subjects: a meta-analysis | + for<br>reduction<br>of total<br>cholesterol<br>(TC), low<br>density<br>lipoprotein<br>(LDL)<br>And<br>unclear for<br>HDL and<br>TG | 2019 | P | 12 | (115) |
| 18 | Improved<br>glucose and<br>lipid<br>metabolism | Probiotics improve<br>glucose and lipid<br>metabolism in<br>pregnant women: a<br>meta-analysis | + | 2019 | T | 13 | (8) |
| 19 | Antioxidant status | Effects of probiotics and synbiotic supplementation on antioxidant status: A meta-analysis of randomized clinical trials | + | 2019 | T | 16 | (116) |
| 20 | Overweight and obese adults | Effects of oral supplementation with probiotics or synbiotics in overweight and obese adults: a systematic review and metaanalyses of randomized trials | + for Waist circumference<br>Effect less for Body weight & BMI | 2019 | P | 28 | (42) |
| 21 | Oxidative stress | The Effects of Supplementation with Probiotic on Biomarkers of Oxidative Stress in Adult Subjects: a Systematic Review and Meta-analysis of Randomized Trials | + | 2019 | T | 11 | (117) |
| 22 | Reduce the weight gain & improve obesity related metabolic parameters | The Potential Role of Probiotics in Controlling Overweight/Obesity and Associated Metabolic Parameters in Adults: A<br>Systematic Review<br>and Meta-Analysis | + | 2019 | T | 7 | (43) |

**Table 3.**
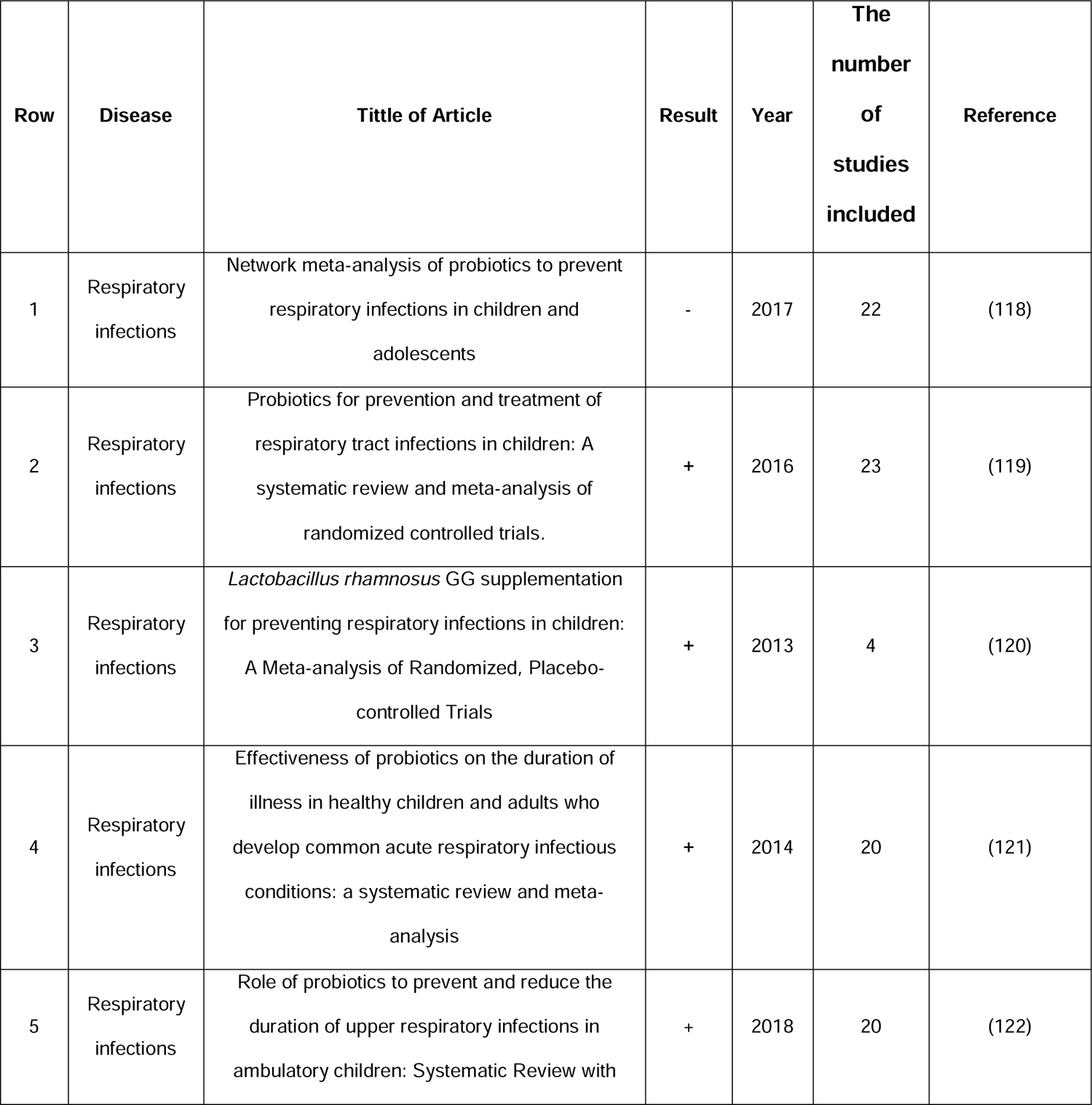
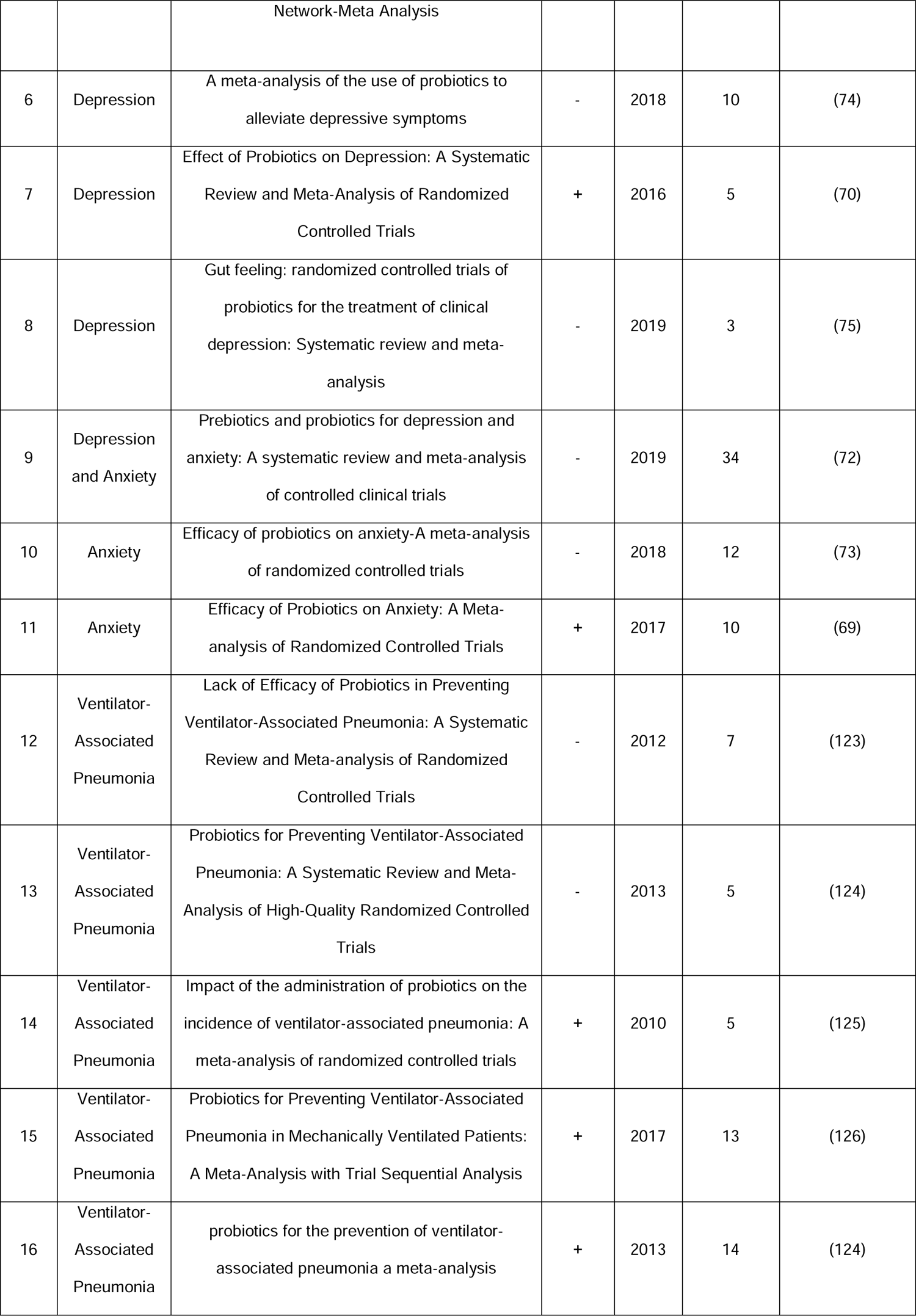
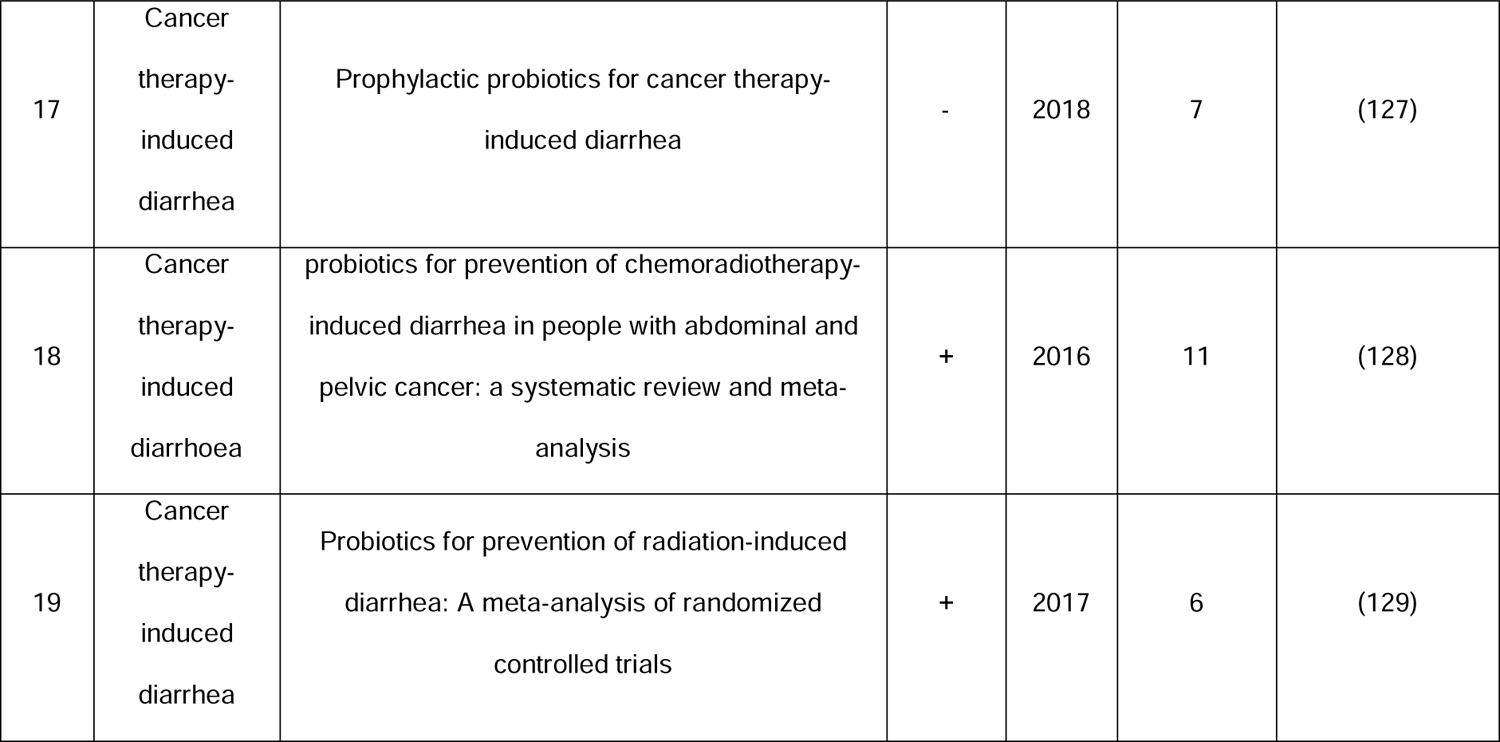
Contradiction reports about the efficacy of probiotics in some diseases Sign + indicates the positive effect of probiotics on the disease. Sign - Indicates that probiotics do not have a positive effect on patients (nor having a negative effect on patients).

Probiotics, which can be effective in decreasing serum cholesterol (hypocholesterolemia), play their role by attaching cholesterol and fatty acids to the membrane of probiotic bacteria(64), deconjugation of bile acids is due to the presence of bile salts hydrolase enzymes (BSH) in lactic acid bacteria(65–68) and conversion of cholesterol to coprostanol and its excretion through the feces(64).

Figure 14 shows the diseases of the genitourinary system. Although various diseases have been studied, the number of studies for each disease is low.

**Figure 14.**
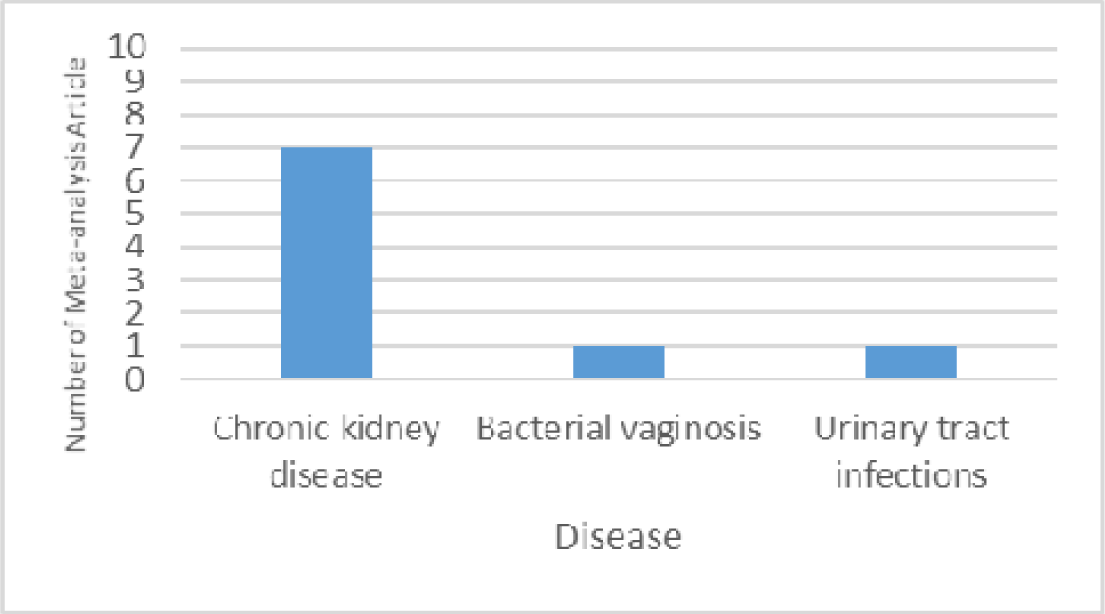
Distribution of meta-analysis articles based on genitourinary system’s disease.

The variety of diseases that have been studied to find the probiotic effect also includes mental illnesses. In Figure 15, the statistics of these cases are illustrated. Depression and anxiety are some of the behavioral and mental disorders that have been studied, although not many studies have been reported, yet half of the studies reported a positive effect of probiotic therapy (69–72) However, half of them did not have any positive or negative effects on these patients(73–76)Recent research suggests that there are two hypotheses through which probiotics affect mood and cognition (77) one of them is regulation of important neurotransmitters and the other is regulation of inflammatory markers by these probiotics (77). Inflammatory factor increases cytokines. Cytokines impact on the synthesis, release and uptake of neurotransmitters(78–80). Finally, the increase in cytokines is associated with neurodegenerative disorders such as depression and anxiety. Probiotics improves parameters such as mood and insomnia in patients with depression (81–83) by reducing inflammatory cytokines Interleukin-1 beta (IL-1) (84, 85), Interleukin-6 (IL-6), tumor necrosis factor-alpha (TNF-α) (81–84).

**Figure 15.**
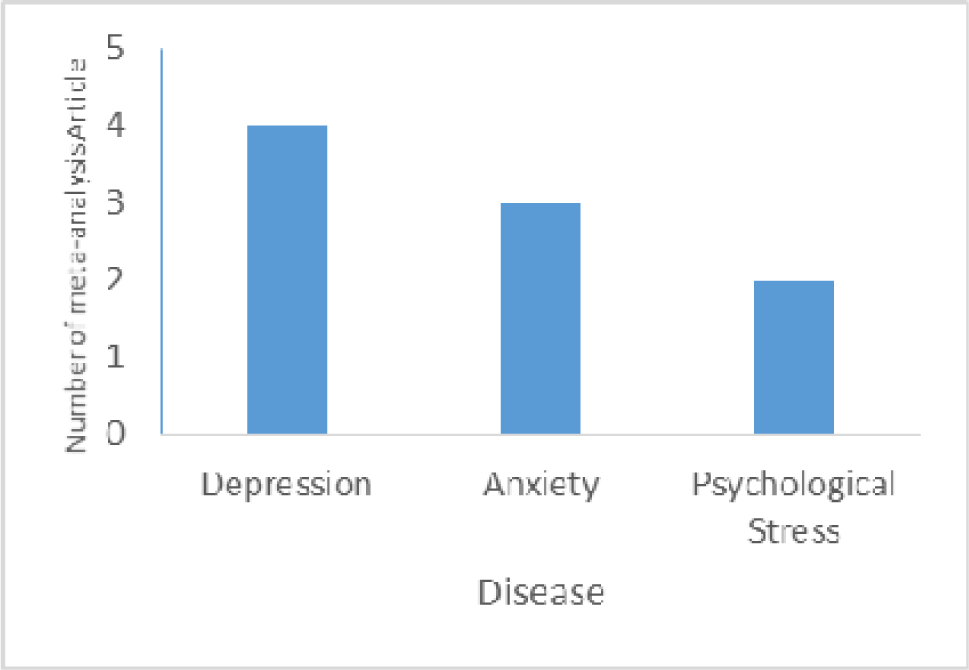
Distribution of meta-analysis articles based on Mental and behavioural’s disorders.

Few studies are also related to injury, poisoning and certain other consequences of external causes, Figure 16.

**Figure 16.**
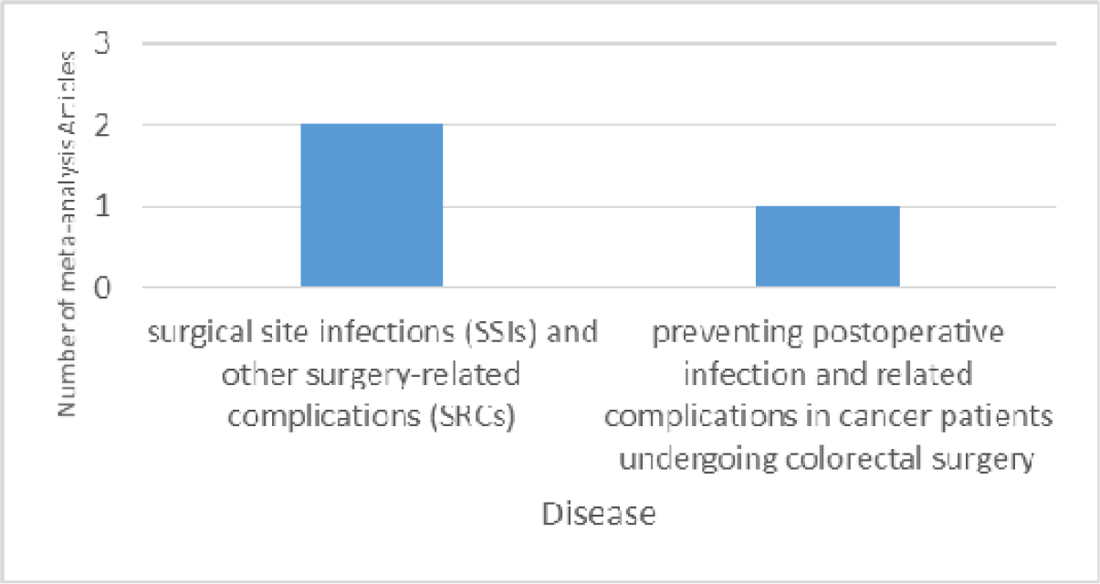
Distribution of meta-analysis articles based on injury, poisoning and certain other consequences of external causes.

Figure 17 shows the statistics of the meta-analysis articles on probiotic effects on other diseases, among which allergies have the highest frequency. According to 6 meta-analysis studies in allergic diseases, the effectiveness of probiotics in preventing and treating this disease is 71.42%. In fact, probiotics reduce allergy symptoms by improving immune function, helping to destroy allergen proteins, confronting pathogenic bacteria, increasing mucus production and strengthen mucosal barrier (86–88). A meta-analysis article on AIDS has investigated the effect of probiotics on CD4 cell count in HIV patients, which reported that probiotic had no effect on CD4 cell count (89) CD4 cell decline is associated with progression of the disease (90, 91). It has been shown that the composition of the gut microbiota changes after body is infected with the HIV, and this change plays a key role in the pathogenesis of HIV (91). Some studies have shown that probiotics play a protective role and also treatment in HIV-infected individuals by modulating immune function (91–93). While other studies have not observed any significant effect of probiotics on the immune system of individuals with this disease(94–96).

**Figure 17.**
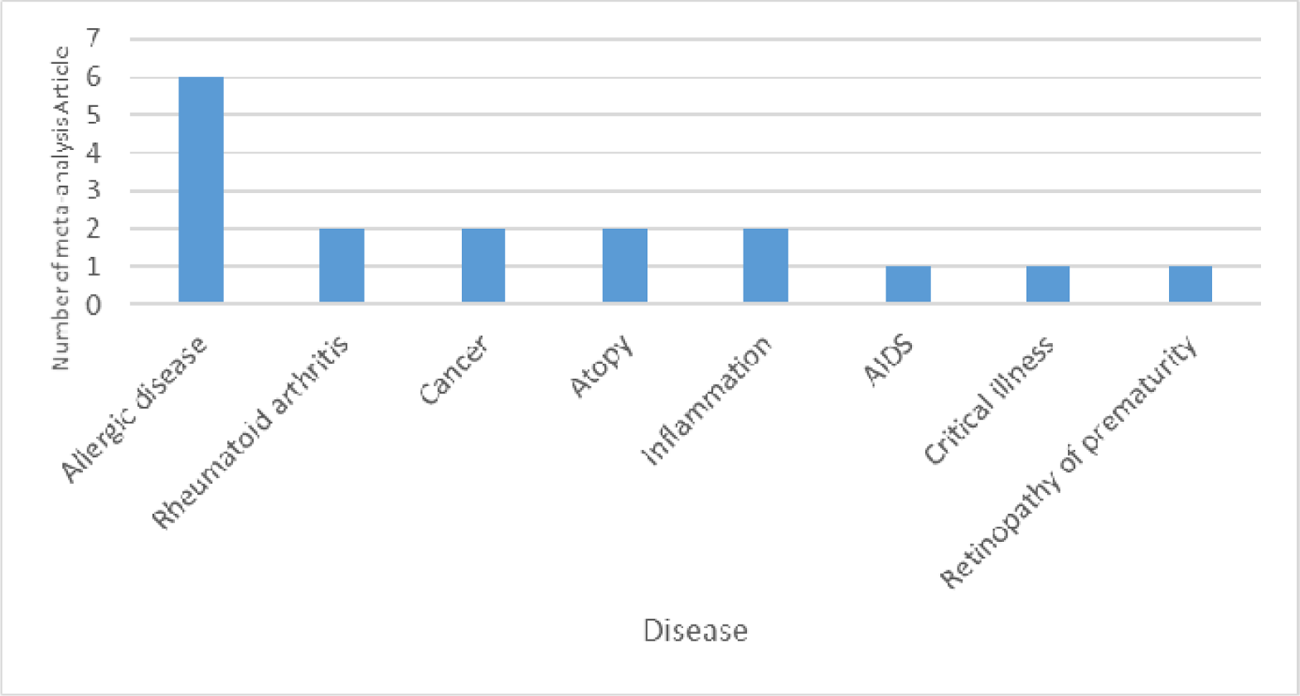
Distribution of probiotics-disease meta-analysis articles based on other disease

Although only two studies have been conducted on cancer, the results suggest that probiotics have been quite effective in preventing cancer. Since half of all cancers are preventable, it is suggested to do further studies on the effect of probiotics in preventing various cancers and its advancing, especially lung, liver, colorectal, gastric and breast cancers, that are the deadliest cancers respectively (97). Among the most important mechanisms that can explain the preventive role of probiotics against the cancer are changes in the gut microbiota, anti-tumor effect through inactivation of cancer compounds, competition with pathogenic microorganisms, improvement the hosts immune response, decomposition of non-digestible compounds, inhibiting tyrosine kinase signaling pathways and antiproliferative effect of cells through regulation of apoptosis and cell differentiation (98–101).

Some articles have not directly examined the effect of probiotics on diseases but have examined the role of probiotics on human health. For example, probiotic effects on pregnancy, oxidative stress or weight gain cannot be categorized as diseases. These articles, totaling 22, are listed in Table 2.

According to the results, probiotics have no effect in improving or preventing retinopathy (1 meta-analysis study including 11 studies), bronchopulmonary dysplasia (1 meta-analysis study including 15 studies) and invasive fungal infections (1 meta-analysis study including 8 studies), rheumatoid arthritis (2 meta-analysis studies including 9 and 4 studies, Acute pancreatitis (4 meta-analysis studies including 4, 6, 7 and 12 studies), asthma (5 meta-analysis studies including 25, 5, 11, 9 and 19 studies) and AIDS (1 meta-analysis study including 11 studies). Although due to the low number of meta-analysis studies on these diseases, may not be *true* to say that probiotics are completely ineffective on these diseases, and further studies are needed to confirm these findings.

On the other hand, probiotics, have been completely effective (100%) on a range of diseases including: *Helicobacter pylori* infection (18 meta-analysis studies include 357 studies), diabetes (15 meta-analysis studies include 264 studies), sepsis (7 meta-analysis studies include 188 studies), ulcerative colitis(7 meta-analysis studies include 143 studies), atopic dermatitis (9 meta-analysis studies include 129 studies), lipid profile (11 meta-analysis studies include 124 studies), hepatic encephalopathy (9 meta-analysis studies include 84 studies), pouchitis (3 meta-analysis studies include 64 studies), gastrointestinal disorder (6 meta-analysis studies include 54 studies), atopy (2 meta-analysis studies include 42 studies), eczema (4 meta-analysis studies include 38 studies), colorectal surgery (3 meta-analysis studies include 36 studies), blood pressure (3 meta-analysis studies include 33 studies), cancer (2 meta-analysis studies include 27 studies), inflammation (2 meta-analysis studies include 23 studies), psychological Stress (2 meta-analysis studies include 19 studies), colic (2 meta-analysis studies include 10 studies), cutaneous wounds (1 meta-analysis study includes 6 studies), cold common (1 meta-analysis study includes 10 studies), critical illness (1 meta-analysis study includes 30 studies), bacterial vaginosis (1 meta-analysis study includes 12 studies), urinary tract infections(1 meta-analysis study includes 5 studies),, intraventricular hemorrhage (1 meta-analysis study includes 32 studies), oral candidiasis (1 meta-analysis study includes 4 studies), pathological neonatal jaundice (1 meta-analysis study includes 13 studies), In the previous section we discussed the salutary effects and mechanisms of probiotics in various diseases.

## Conclusion

The study of the probiotic effect in the prevention or treatment of diseases has long attracted the attention of many researchers. In recent years these studies have increased significantly in a way that so far from 2000 to October 2019, 283 articles on the probiotic effect in the prevention and treatment of various diseases have been reported directly and 22 cases have been reported indirectly, role review of probiotics have been reported on human health and not directly on disease. A comprehensive systematic review of meta-analyses has several benefits in understanding the distribution and developing attitudes towards probiotics in different diseases and orienting the future research. Creating a perspective from research in this area may open new horizons for researchers, thus make them capable to initiate wiser projects. Most significant features of this study can be briefly noted below.

The increasing trend of publication of articles by year indicates the growing interest of researchers in the field, which has recently become one of the most attractive subjects for researchers interested in probiotics. There is an actual starting point of consideration of meta-analyses research for probiotics form 2011. In 2018, published articles frequency is at its peak. Since the studies have been reviewed until October 2019, given the ascending trend, maybe taking into the account the second half of the 2019 year will lead to a higher peak. Therefore, growing in number of articles is expected to continue in the coming years. Some currently accepted papers from before, may be under publication, and will appear in near future, also.

According to the results of this study, among the most meta-analysis studies (diseases with more than 10 original studies), probiotics were more than 90% effective in preventing and treating diseases. Some articles have reported no positive effects in disease incidence or preventing its progression thus make it essential to perform further study to confirm this. Since some meta-analysis articles suggest no effect while other suggests a positive effect for the same disease, it might be interesting for future works to see if there is a correlation between the number of studies in a meta-analysis and the outcome (effective or not effective). Our investigation on disease having more than 10 meta-analyses research (total 170 study including 519622 participants) shows that the average effectiveness of probiotics is more than 90% for the diseases with top most meta-analysis count (Table 4).

**Table 4.** Relativity of the nember of the meta-analysis studies with the effectiveness report of probiotics on the dieseaes.

| Disease | Meta-analysis count | Total number of the studies in the meta-analyses | Number of subjects | Effectiveness percent |
| --- | --- | --- | --- | --- |
| Infections | 47 | 874 | 150310 | 93.61% |
| Diarrhea | 40 | 669 | 120306 | 80.95% |
| Preterm Infants | 23 | 467 | 110984 | 82.60% |
| Enterocolitis | 20 | 395 | 91155 | 85% |
| Diabetes | 15 | 264 | 15966 | 100% |
| Irritable bowel<br>syndrome | 14 | 236 | 18952 | 92.85% |
| Lipid profile | 11 | 124 | 11949 | 100 % |
| Total | 170 | 3029 | 519622 | 90.71% |

However, none of the investigated papers has reported the detrimental effect of probiotics on human health. Further studies are needed to confirm the effectiveness of probiotics in the treatment, prevention or improvement of diseases, especially to confirm the effectiveness of probiotics in less studied diseases (such as AIDS, cancer, ocular and musculoskeletal diseases). For the AIDS and cancer, due to the weakness of immune system, patients are at risk for secondary infections. Therefore, further investigation of the role of probiotics in the prevention or treatment of subsequent infections may be important. However, there is strong evidence that probiotics play a beneficial and effective role in the prevention and treatment of several diseases including: diarrhea, infection, necrotizing enterocolitis, irritable bowel syndrome, type 2 diabetes, *Helicobacter pylori* eradication and necrotizing enterocolitis, sepsis in premature infants and lipid profile. The scope of studies in this article can also be expanded to Cochrane reviews. Our study is only limited to probiotics related meta-analysis studies, and it cannot be generalized to other studies about probiotics. There are rare negative outcomes related to probiotics such as (11) that indicates using probiotics for severe acute pancreatitis (PROPATRIA) did not get better results, but it was associated with an increased risk of mortality. It should be mentioned that since the writing of this article (from September 2018 to the October 2019), 49 other relevant meta-analysis studies have been published which according to our prediction this upward trend in the number of meta-analysis articles that scrutinize the effect of probiotic on various diseases will continues to increase. For the future works, Cochrane reviews, meta-analysis including dozens of articles (as e.g. for NEC and AAD) may be included in the study and the quality of the articles can be considered.

## Data Availability

Not applicable. All the used data is available in the paper references.

## Acknowledgments

It’s pleasure to express our deep sense of gratitude to professor Arthur C. Ouwehand from DuPont Nutrition and Biosciences for his valuable and directive critical reviews and comments of this research.

## Statement of Competing Interests

The authors have no competing interests.

## List of Abbreviations

R: Result

P: Prevention

T: Treatment

BMI: Body mass index

ICD: International Classification of Diseases system

CD: Crohn’s disease

UC: Ulcerative colitis

SCFAs: Short chain fatty acids

GABA: Gamma amino butyric acid

BSH: Bile salts hydrolase enzymes

IL-1: Interleukin-1 beta

IL-6: Interleukin-6

TNF-α: Tumor necrosis factor-alpha

TC: Total cholesterol

LDL: Low density lip^β^oprotein

## Notes

### Competing Interest Statement

The authors have declared no competing interest.

