## Supplemental Figure S.1 and Table S.1 for "Twenty years review of probiotic meta-analyses articles: Effects on disease prevention and treatment"

**Supplementary Materials:**

Number of Articles

| Total Number of Articles per Year | Number of Articles |
| --- | --- |
| 2000 | 1 |
| 2001 | 0 |
| 2002 | 4 |
| 2003 | 1 |
| 2004 | 0 |
| 2005 | 1 |
| 2006 | 8 |
| 2007 | 3 |
| 2008 | 10 |
| 2009 | 6 |
| 2010 | 9 |
| 2011 | 5 |
| 2012 | 12 |
| 2013 | 18 |
| 2014 | 16 |
| 2015 | 28 |
| 2016 | 31 |
| 2017 | 47 |
| 2018 | 59 |
| 2019 | 20 |

Fig. S.1 Distribution of meta-analyses on the effect of probiotics on disease risk and disease management by year till June 2019

Table S.1 The top 15 journals publishing most probiotics-diseases related meta-analysis articles till June 2020

| Journal | Number of Article |
| --- | --- |
| Alimentary Pharmacology & Therapeutics | 16 |
| PLOS ONE | 11 |
| Medicine | 9 |
| Pediatrics | 7 |
| World Journal of Gastroenterology | 7 |
| Nutrients | 6 |
| Digestive Diseases and Sciences | 5 |
| Gastroenterology | 5 |
| British Journal of Nutrition | 4 |
| Acta Paediatrica | 3 |
| Annals of Allergy, Asthma & Immunology | 3 |
| Clinical Nutrition | 3 |
| Clinics and Research in Hepatology and Gastroenterology | 3 |
| Critical Care | 3 |
| Critical Care Medicine | 3 |
